# Barrier gesture relaxation during vaccination campaign in France: modelling impact of waning immunity

**DOI:** 10.1101/2021.08.29.21262788

**Authors:** Carole Vignals, David W. Dick, Rodolphe Thiébaut, Linda Wittkop, Mélanie Prague, Jane Heffernan

## Abstract

Non-pharmaceutical interventions have been implemented intermittently for more than a year in most countries of the world to mitigate COVID-19 epidemic. In France, while the vaccination campaign is progressing, the French government has decided to remove many public health restrictions such as business closure, lockdowns and curfews. Nonetheless, social distancing, mask wearing, and hand washing (also called barrier gestures) are still recommended. We utilize an age-structured compartmental SEIR model that takes into account SARS-CoV-2 waning immunity, vaccination, and increased transmissibility from variants of concern, to estimate if barrier gestures can be relaxed without causing a resurgence of severe infections. This model assumes that susceptibility to infection is a function of immunity status, which depends on initial infection severity and vaccination status. It is calibrated on confirmed COVID-19 cases from the French surveillance database, and accounts for changes in contact behaviors due to implementation of nation-wide public health policies. We study partial and full relaxation of barrier gestures occurring from August to December 2021 under various immunity duration assumptions. Maintaining application of barrier gestures appears essential to avoid a resurgence of severe infections that would exceed health care capacities, while surmounting vaccine hesitancy represents the key to consider their relaxation. Immunity duration assumptions significantly influence the short-term dynamic of the epidemic which should be considered for further modelling.

## 1. Introduction

Causing more than 200 million infections and at least 4.4 million deaths worldwide, the COVID-19 epidemic is a global health, economic, and social crisis. The causative agent, SARS-CoV-2, emerged in China’s Hubei province in December 2019. Various non-pharmaceutical measures, in particular lockdown and social distancing, have been implemented around the world to effectively control the spread of the virus ^1 2^. Nevertheless, all of these measures have negative economic and social consequences, as well as repercussions on the mental health of populations. Considerable efforts in vaccine research have resulted in the development of several effective vaccines against COVID-19 in a few months. As of August 4^th^, 2021, more than 3,8 billion doses of vaccine have been administered worldwide ^3^.

Numerous studies have used compartmental models to either estimate the parameters of the epidemic dynamics ^4 5^, measure the impact of social distancing ^6 7 8^ or evaluate different vaccine strategies ^9^. Many of these studies assumed that immunity acquired after an infection was definitive. Indeed, it has been shown that immunity acquired after a first infection protects against reinfection for at least 6 months ^10 11^. Nevertheless, several immunological studies have shown that, following natural infection, the anti-SARS-CoV2 antibody titer declines over time ^12 13 14 15^ even though it remains detectable in more than 90% of cases at 7 months ^14^. A study suggests that anti-SARS-CoV-2 CD4+ and CD8+ specific T cells decline with a half-life of 3 to 5 months ^15^, while another study found that they were maintained 10 months post-infection ^16^. With regard to other coronaviruses, reinfections are frequent in the year following a seasonal coronavirus infection (common cold) ^17^, while immunity appears to be prolonged after infection with SARS-CoV or MERS ^18 19 20^. Concerning vaccination, a significant trend of anti-SARS-CoV2 antibodies declining have been demonstrated with both BNT162b2 and ChAdOx1 nCoV-19 ^21^. The latest study endpoint of the BNT162b2 clinical trial suggests that vaccine efficacy against symptomatic infections may decline over time ^22^. However, to date the exact duration of the protection anti-SARS-CoV-2 conferred by an infection or by vaccination remains unknown. As the epidemic has lasted for more than a year, it seems necessary to take this parameter into account to modelling the epidemic. Indeed, several studies have shown that the future evolution of the epidemic would depend on the duration of immunity, from extinction after a large epidemic wave to the transition to an endemic mode ^23 24 25 26^.

The French government, as many other, has successively implemented many restrictions in order to mitigate the epidemic, such as travel restrictions, lockdowns, curfews, obligation to work from home, closure of bars, restaurants, cinemas or schools. In addition to these measures, barrier gestures have been promoted. These barrier gestures include mask wearing, washing hands regularly, using a single-use disposable tissues, keeping a distance of at least 2 meters between individuals and avoiding gatherings of more than 6 peoples, with the aim to reducing human-to-human viral transmission.

Four vaccines are currently licensed in France, two messenger RNA vaccines (BNT162b2 and mRNA-1273), and two viral vector vaccines (ChAdOx1 nCoV-19 and Ad26.COV2.S). The vaccination campaign began on January 2021, targeting first the elderly and the subpopulation with comorbidities at risk for severe COVID-19. The program was then progressively extended to entire adult population over a few months, and then to children over 12 years old. As of August 17^th^, 2021, 69.5% of the French population have received at least one dose of vaccine and 59.3% have completed the vaccination schedule ^27^.

A French modelling study estimated that it would be necessary to vaccinate 90% of individuals over 65 years old and 89% of the 18-64 year old’s before all social distancing measures could be relaxed ^28^. This study assumes that immunity does not wane over time, which could lead to an underestimation of these thresholds. In addition, Europe is facing the breakthrough of the delta variant of concern, which has been estimated to be 1.97 times more transmissible than the historical strain, and is predicted to represent 90% of COVID-19 cases in August 2021 ^29^. However, the vaccine coverage reached at the end of the vaccination campaign is uncertain. A survey conducted among the French population estimates that 16% of adults do not want to be vaccinated and that only 74% of parents would agree to have their child under 17 vaccinated ^30^. Finally, because immune responses following vaccination are lower in the elderly ^31^, a booster campaign for this population is being considered by the authorities.

While the vaccine rollout is progressing, the French government has removed public health restrictions, but the application of barrier gestures is recommended ^32^. In this study, we aim to estimate the consequences of a barrier gesture relaxation on the evolution of the epidemic in France, taking into account vaccination, the emergence of variants of concern, in particular, the delta variant, and waning immunity. In addition, we aim to investigate in a preliminary analysis the impact of a booster vaccine campaign for the elderly.

## 2. Materials and Methods

### 2.1. Model

We used a compartmental age-structured model of COVID-19 infection previously developed by Childs et al. ^33^ which is adapted from a model of vaccination and waning immunity applied to pertussis ^34^. A flow diagram of the model is shown in **Figure 1** for a single age group. The model is based on a Susceptible-Exposed-Infected-Vaccinated-Susceptible model structure (SEIVS) with age structure (i.e., groups 0-4, 5-9, …, 75+ years). As in Childs et al. ^33^, we use S_im_, E^k^_jm_, I_jm_, and V^l^_im_ to denote the number of susceptible, exposed, infectious and vaccinated individuals in each age group m (1 ≤ m ≤ 16), where i (1 ≤ i ≤ 4) denotes immune status, j (2 ≤ j ≤ 4) denotes symptom severity, k (1 ≤ k ≤ 3) represents stages in the exposed class, and l (1 ≤ l ≤ 2) denotes the number of vaccine doses that individuals have received.

**Figure 1.**
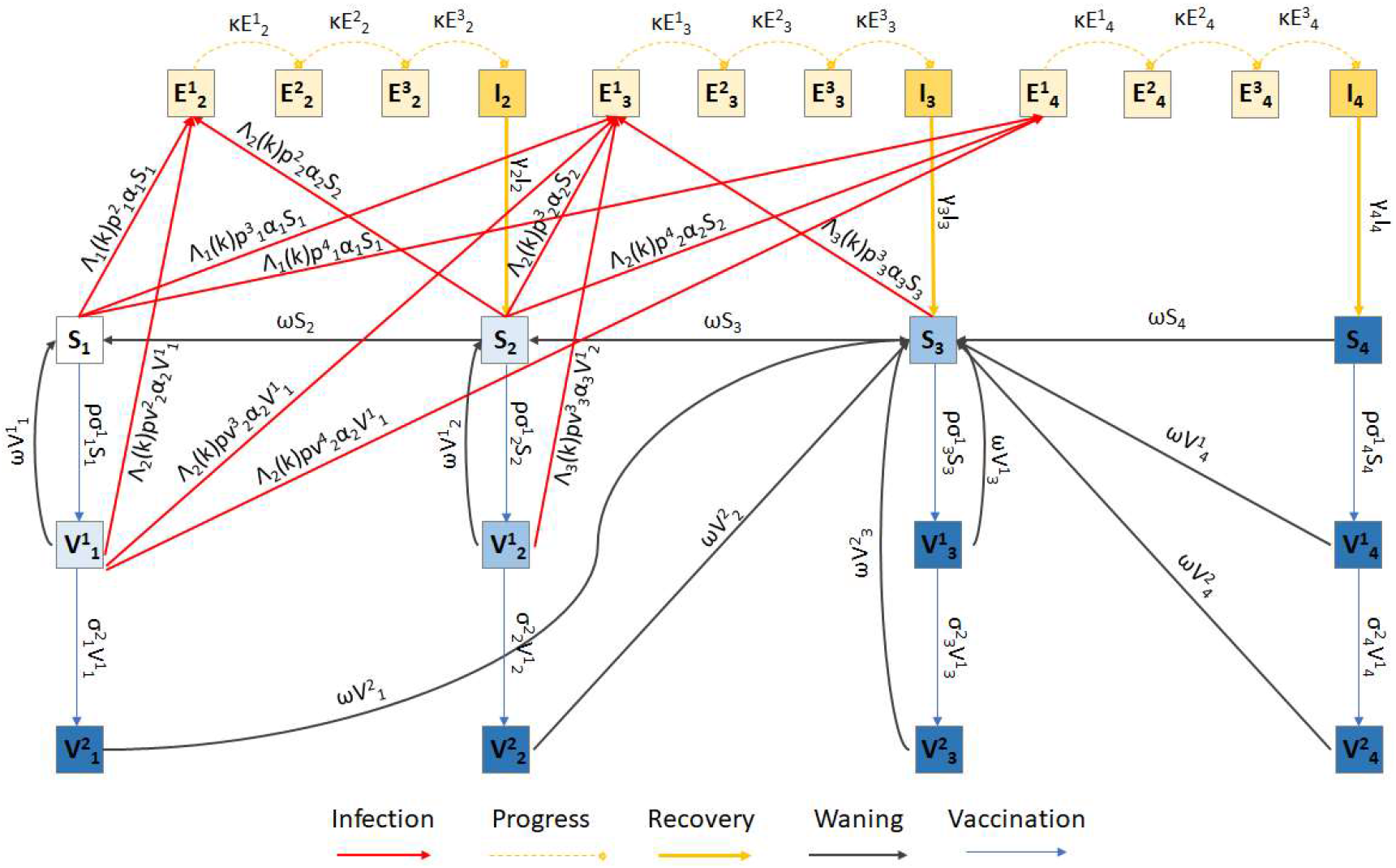
Schematic of the age-structured SEIVS model for one age group, derived from Childs et al.

A full description of the model is available in Childs et al. ^33^. Briefly, the base model consists of an immune continuum, distinguishing four states of susceptibility (fully susceptible (S_1_), somewhat immune (S_2_), moderately immune (S_3_), and fully resistant to infection (S_4_)), three infectious states with mild (I_2_), moderate (I_3_), and severe (I_4_) symptoms (i.e., infections requiring at least one medical consultation), and three infected but not-yet-infectious states (E^k^_j_, j= 2,3,4, k= 1,2,3). It is assumed that individuals of higher immune status are less susceptible to infection than those of lower status. Co-morbidity statuses by age ^35^ determine the probability of mild, moderate, and severe symptoms for each age group. Immunity develops after infection. It is assumed that people with mild, moderate, and severe symptoms move to immune classes S_2_, S_3_ and S_4_, respectively, upon recovery (i.e., the severity of symptoms is proportional to neutralizing immunity development ^14 36 37 38^). Vaccination is implemented into the model using a two-dose structure, where compartments V^1^_i_ and V^2^_i_ denote those with one and two doses, respectively, where *i* is the level of susceptibility. It is assumed that two doses of vaccine, or one dose of vaccine administered to individuals in immune states S_3_ and S_4_, provide the same level of immunity or protection from infection as S_4_; i.e., fully resistance to infection. Additionally, it is assumed that one dose of vaccine given to individuals in S_1_ and S_2_ provides protection similar to states S_2_ and S_3_, respectively, where protective efficacies against infection and/or severe disease decrease with a lower level of immunity. Finally, it is assumed that immunity gained from infection or vaccination wanes over time (**Figure 1**, black lines).

Detailed descriptions of the equations are provided in appendix A.

### 2.2. Parameters

Parameter values and references are provided in **Table 1**.

**Table 1.** Parameter definitions, values and references for the age-structured SEIVS model

| Parameter | Definition | Value |  |  | References |
| --- | --- | --- | --- | --- | --- |
| Fixed parameters |  |  |  |  |  |
| $\alpha_i$ | Susceptibility by immune status i | $\alpha_1$ | $\alpha_2$ | $\alpha_3$ | Hypothesis, taken from Childs et al. <sup>33</sup> |
|  |  | 1 | 0.66 | 0.33 |  |
| $\beta_j$ | Infectivity by infection severity j | $\beta_2$ | $\beta_3$ | $\beta_4$ | <sup>39</sup> |
|  |  | 0.045 | 0.089 | 0.009 |  |
| $\lambda_{im}$ | Force of infection by immune status i and age m | Appendix A – Model equations | | | Estimated |
| $c_{ma}$ | Contact-rates between individuals in age group m and age group a | Described in Section 2.3 | | | <sup>40</sup> |
| R0 | Basic reproductive number | 2.9 |  |  | <sup>4</sup> |
| $p_{im}^j$ | Proportion of S going to I<sub>j</sub> by age m and by immune status i | Table S2 | | | <sup>35</sup> |
| $A_m$ | Population size by age m | Table S1 | | | INSEE |
| $\gamma_j$ | Recovery rate by infection severity j | $\gamma_2$ | $\gamma_3$ | $\gamma_4$ | Hypothesis |
|  |  | 1/5 | 1/10 | 1/15 |  |
| $\kappa$ | Rate of progress through the exposed compartments | 1/1.5 | | | <sup>41 42 43</sup> |
| Variable parameters |  |  |  |  |  |
| $\sigma^1_{im}$ | Vaccination rate by age m and by immune status i for first dose | Described in Section 2.2 paragraph 2 | | | VAC-SI* |
| $\sigma^2_{im}$ | Vaccination rate by age m and by immune status i for second dose | 1/28 | | | |
| $\omega$ | Waning rate of immunity | Main analysis | Sensitivity analysis | | <sup>21 22</sup> |
|  |  | 1/1095 | 1/365 | 0 |  |
| $\rho$ | Vaccine efficacy against infections after 2 doses | Main analysis | Sensitivity analysis | | <sup>44 45 46 47</sup> |
|  |  | 0.8 | 0.9 |  |  |
| 1 - $\varepsilon$ | Vaccine efficacy against infections after 1 dose | Main analysis | Sensitivity analysis | | <sup>44 45 46 47</sup> |
|  |  | 0.5 | 0.7 |  |  |
| 1 - q | Vaccine efficacy against severe cases after 1 dose | Main analysis | Sensitivity analysis |  | <sup>44 45 46 47</sup> |
|  |  | 0.7 | 0.7 |  |  |

\*VAC-SI = Système d'Informations pour le suivi de la VACCination/ INSEE = *Institut National de la Statistique et des Etudes Economiques*

Susceptibility is assumed to decrease with increasing immunity, but does not depend on age. Thus, susceptibility α of an individual somewhat immune (S_2_) or moderately immune (S_3_) correspond respectively to 1/3 and 2/3 of that of an individual fully susceptible (S_1_). Infectivity (β) is assumed to vary by severity of disease and is chosen to produce a basic reproduction number, R0, equal to 2.9 using the Next Generation Matrix method ^4 48^. By immunity status, we assume that the infectivity of individuals with mild (I_2_) or severe (I_4_) infections is 0.5 and 0.1 times the infectivity of moderate infections (I_3_). Indeed, people with milder symptoms are expected to have lower infectivity ^39^. Simultaneously, more severe disease outcomes are expected to induce behavioural changes, such as limiting mobility, that lower infectivity. Infectivity is considered in the calculation of the infection force λ, which takes into account the average number of contacts between age classes and the proportion of the population infected and infectious (Appendix A – Model equations). Susceptible individuals S_i_, upon infection, move to the mild, moderate, and severe symptom classes with probabilities p^j^_im_ for each age group m and each immune status. These probabilities are determined by the prevalence of zero, one, and two or more co-morbidities that increase the risk of severe COVID-19 disease within each age group, informed by ^35^. p^j^_1m_ is directly derived from these prevalences, whereas p^j^_2m_ and p^j^_3m_ are modified in order to assume that people in the S_2_ and S_3_ compartments are, respectively, 70% protected and fully protected against moving to the severe disease class I_4_ (Table S2). The incubation period lasts on average 4.5 days, therefore, it is assumed that the progression rates through the pre-infectious period κ is equal to 1/1.5. The contagious period is estimated to last 7 to 8 days ^41 49 50^ and depends on symptom severity, with milder disease associated with shorter infectious periods ^51 52^. Thus, we assumed that the recovery rate is respectively equal to 1/5, 1/10 and 1/15 for mild (I_2_), moderate (I_3_) and severe infections (I_4_). Given the short period considered, we assume the absence of natality, mortality and aging. Finally, we assume that immunity lasts on average 3 years between successive stages, which means that it takes 9 years to pass from a fully resistant to a fully susceptible compartment. For vaccinated classes, we assume that immunity in V^1^_1_ wanes to S_1_, that in V^1^_2_ wanes to S_2_, and that of all other vaccinated classes wanes to S_3_.

Vaccination is implemented from 01/01/2021 and takes into account the different age groups that are successively eligible in France, starting from the elderly first (Table S3). In order to take into account vaccine hesitancy, we assume that 10% of the over-75s, 20% of the [20-74] years old and 30% of the [10-19] years old will not be vaccinated ^30^. We determine the first dose vaccination rate σ^1^_i_ given the desired coverage at the end of each month of the vaccination program. From January to June 2021, the monthly coverage is derived from the VAC-SI (Système d’Informations pour le suivi de la VACcination) database (Table S4). From July, we assume that 15 million doses (including first and second doses) will be administrated each month, which corresponds to the average distribution in the previous 3 months. Monthly coverage is scaled to a daily rate accounting for the portion of the population in the current vaccine-eligible age groups in each eligible compartment (S_1_, S_2_, S_3_ and S_4_). Individuals receive a second dose 28 days after the first one and acquire immunity immediately. Vaccine efficacy against all infections is assumed to be 50% after 1 dose and 80% after 2 doses. Vaccine efficacy against severe infections is assumed to be 70% after 1 dose. Given the model architecture, the efficacy against severe infections after 2 doses is equal to the efficacy against all infections after 2 doses.

In order to take into account the emergence of the variants of concern (alpha, beta, gamma and delta), we use the SI-DEP database (Système d’Informations de DEPistage), which provides information about the distribution of these variants among all confirmed COVID-19 cases over time (Figures S1 and S2). For the delta variant, we assume that it will represent 90% of the reported cases at the end of August 2021 ^29^. We assume that alpha-beta-gamma variants and the delta variant are, respectively, 50% ^53 54^ and 97% ^55^ more transmissible than the historical strain.

### 2.3. Contact matrices and public health mitigation strategies

Age-specific daily contacts for the French population are obtained from pre-pandemic estimations ^40^. The model incorporates in the contact-rates, c_ma_, the governmental measures taken to mitigate the epidemic, such as lockdowns, curfews, closing schools, business, bars and restaurants or work from home. The schedule of these governmental measures is available at https://www.gouvernement.fr/info-coronavirus/les-actions-du-gouvernement. Specific modifications to the contact matrices are provided in the Supplementary Material (Tables S5 to S8).

### 2.4. Calibration

The model is calibrated using daily reported COVID-19 cases from the SI-DEP database, from 13/05/2020 to 01/07/2021. SI-DEP is the French surveillance system that gathers all COVID-19 infections confirmed by Polymerase Chain Reaction or antigenic test at the national level. We assume that reported COVID-19 cases correspond to all severe cases (I_4_) and 3/5ths of the moderate cases (I_3_). The k-value is the only model parameter that is fit to COVID-19 data and reflects the population compliance to barrier gestures. Given that climatic conditions are not implemented in the model, the k-value also captures part of the influence of the climate on the transmission ^56^. This parameter lies between 0 and 1 (1 being pre-pandemic value; i.e., no barrier gesture application), and it linearly scales contact-rates from the contact matrices. Thus, the force of infection λ_im_ depends on the k-value (Appendix A).

### 2.5. Barrier gesture relaxation scenarios

After August 1^st^, 2021, we formulate different hypotheses for the relaxation of barrier gestures, based on the evolution of the k-value. The baseline scenario corresponds to no relaxation. For this scenario, we chose to set the k-value for July-August 2021 to those estimated during the same period in the previous year, assuming that barrier gestures will be applied at the same level. After August 2021, we maintain the k-value at this level, until it is modified to reflect a relaxation scenario.

Different scenarios of barrier gesture relaxation are formulated according to the timing of the relaxation (August, September, October, November or December) and its magnitude (k-value raised to 0.7, 0.8, 0.9 or 1, i.e., the pre-pandemic level). For each relaxation scenario, the k-value set before the relaxation is identical to the no relaxation scenario. We chose to evaluate the impact of each relaxation scenario on the evolution of severe cases I_4_, i.e., infections requiring at least one medical consultation, and on the number of prevalent intensive care units’ (ICU) hospitalizations. Because our model does not directly incorporate a hospitalized compartment, we extrapolated the predictive number of ICU hospitalizations by estimating the mean ratio between the past ICU hospitalizations from the SI-VIC database (Système d’Information pour le suivi des VICtimes d’attentats et de situations sanitaires exceptionnelles) and the past I_4_ cases.

### 2.6. Sensitivity analyses

We successively explored a pessimistic immunity duration hypothesis, assuming that immunity wanes totally over 3 years instead of 9, and an optimistic hypothesis assuming that immunity does not wane. We also study one scenario without vaccine hesitancy, thus allowing vaccination of 100% of each eligible age class, and one analysis with a better vaccine efficacy (**Table 1**). The model was refit to estimate the retrospective k-value for all of these analyses except for the no vaccine hesitancy hypothesis.

Finally, we implemented (crudely) a booster vaccination campaign for people over 75 in order to estimate the upper bound effect of this strategy. In this analysis, we assumed that, on September 1^st^, 2021, a fraction of people over 75 moves from the S compartments to the corresponding V^2^ compartments, ultimately returning all 75+ individuals that were previously protected by vaccination back to the V^2^ protective classes.

## 3. Results

### 3.1. Calibration

**Figures 2(a)** and **3** show the k-value (reflecting the population compliance to barrier gestures) estimation and the model fit from May 13^th^, 2020 to July 1^st^, 2021 for the main analysis, respectively. Figures showing the k-value and the fit of the model to the data for the sensitivity analyses are shown in the supplementary material (Figures S3, S4 and S5). In addition, **Figure 2(b)** plots the reduction in the average number of contacts for the entire population over time, given the combined effect of the modifications in the contact matrices and the fitted k-value. We note qualitative similarities between this figure and the time-dependent transmission rate of COVID-19 in the French population ^8^.

**Figure 2.**
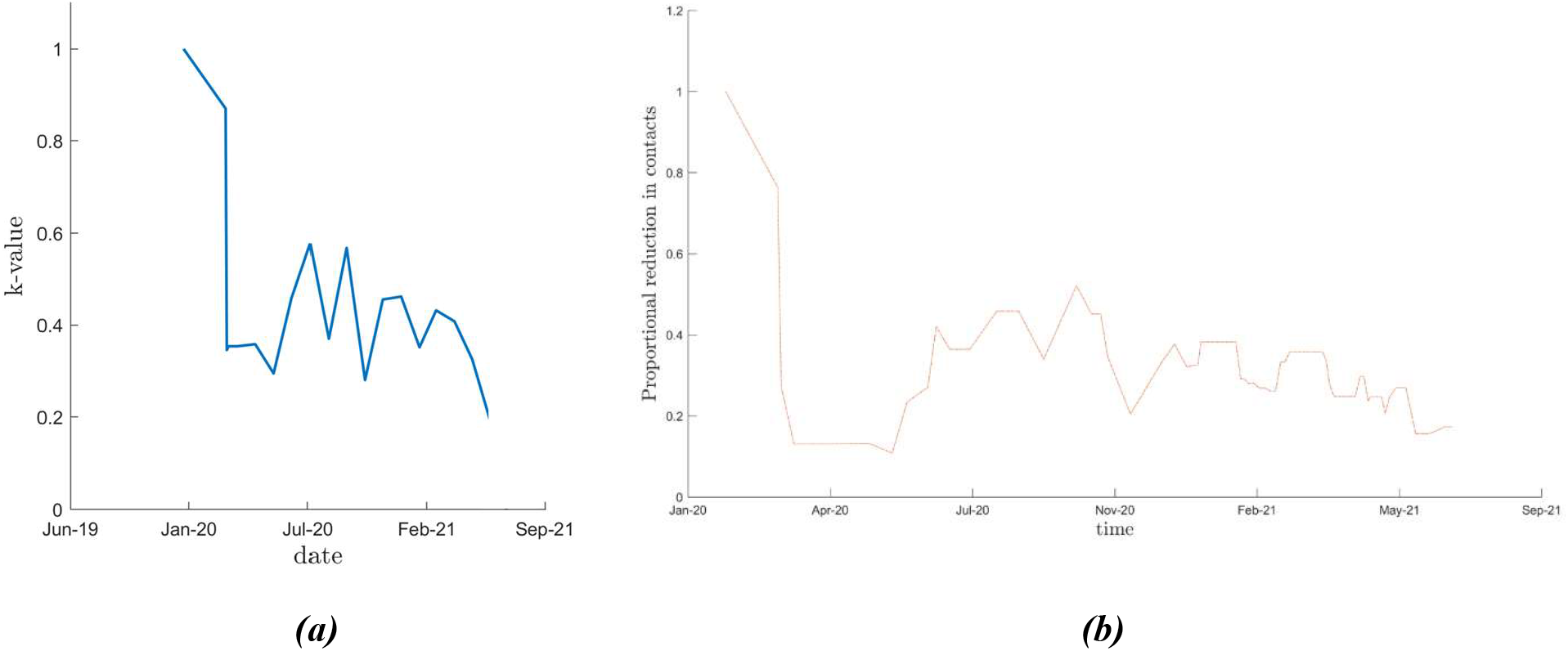
**(a)** Main analysis k-value estimation from May 13, 2020 to July 1^st^, 2021. **(b)** Main analysis estimation of reduction in the average number of contacts consecutively to non-pharmaceutical interventions implemented in France from May 13^th^, 2020 to July 1^st^, 2021

**Figure 3.**
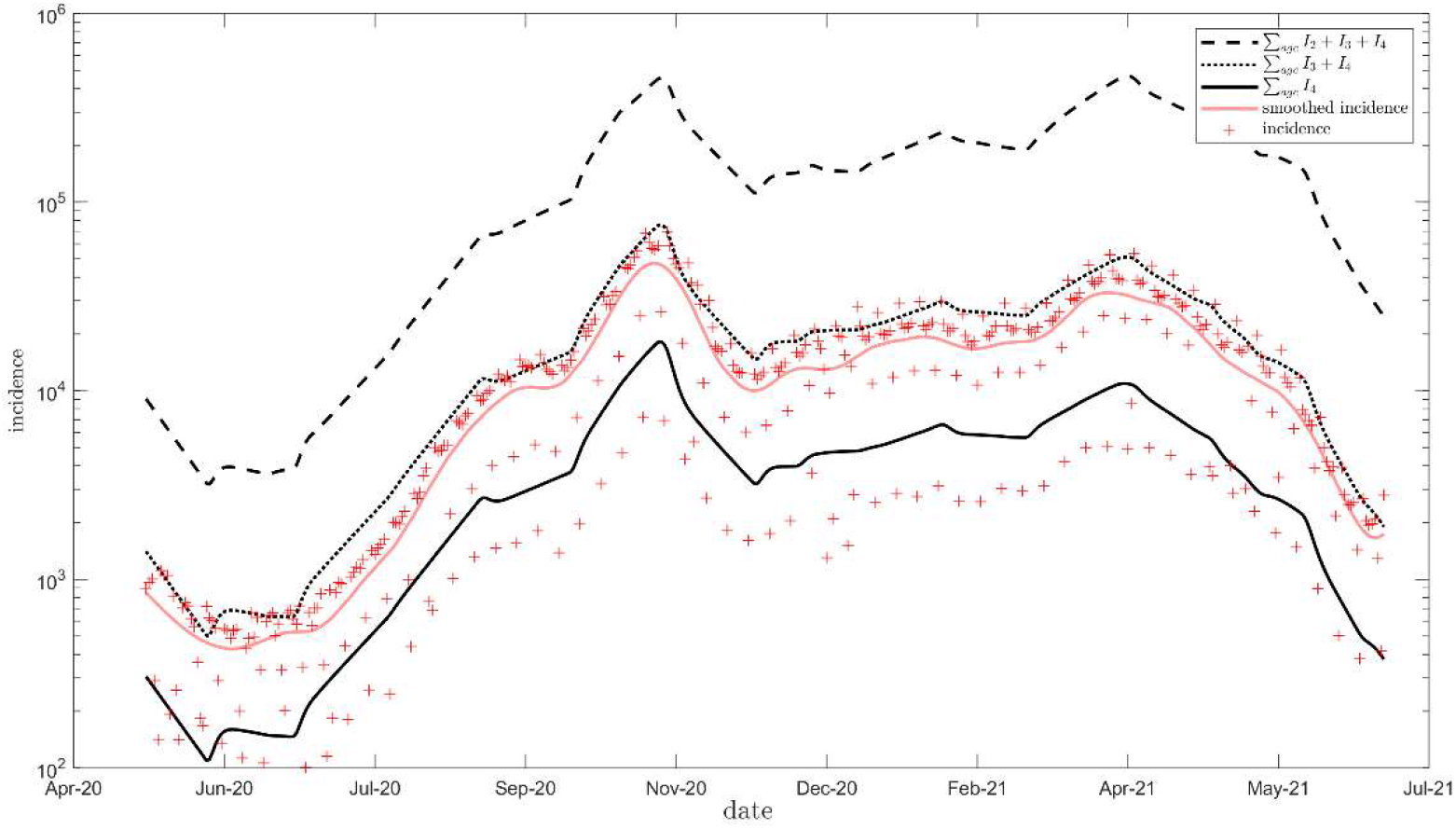
Main analysis model fit (logarithmic scale) from May 13^th^, 2020 to July 1^st^, 2021 to reported COVID-19 cases from French surveillance database (SI-DEP-Système d’Informations de DEPistage)

### 3.2. Impact of barrier gesture relaxation, main analysis

The predicted evolution of severe case I_4_ incidence for each relaxation scenario are presented in **Figure 4**, from July 1^st^ 2021. Regardless of the timing, the complete relaxation of barrier gestures (setting k-value to 1, pink curve) is followed by an epidemic resurgence whose peak systematically exceeds that of the 2^nd^ and 3^rd^ epidemic waves in France. However, even in the optimistic scenario of no relaxation (defining as a barrier gesture applying at an equivalent level to that of the same period in 2020), we do observe a significant rebound (black curve).

**Figure 4.**
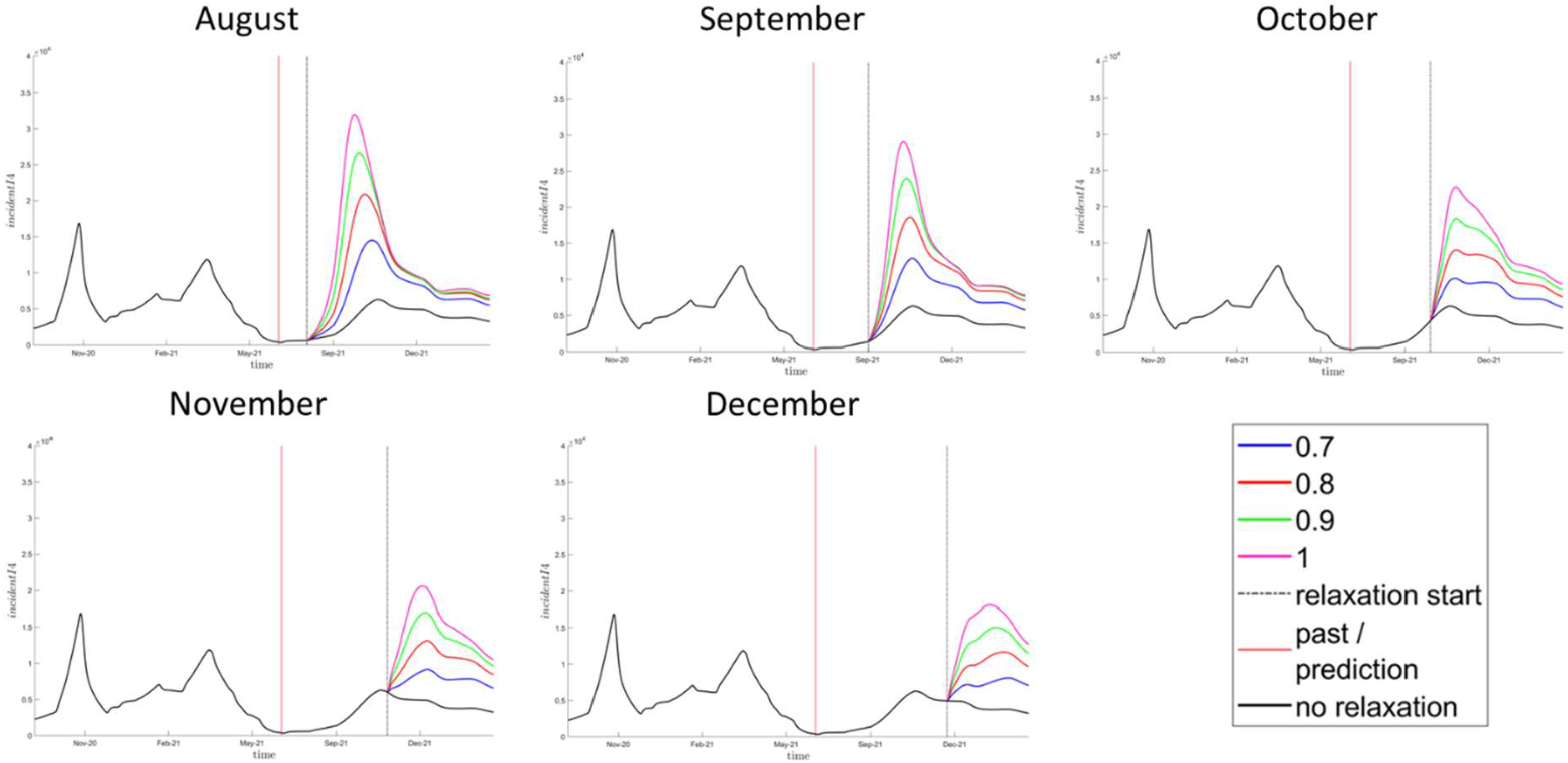
Evolution of incident COVID-19 severe cases (I_4_) in France for each barrier gesture relaxation scenario in the main analysis (i.e., 9 years waning immunity assumption). Relaxation scenarios: k-value (barrier gesture compliance index) raised either to 0.7, 0.8, 0.9 or 1 in August, September, October, November or December. Relaxation’s start is indicated by the vertical dashed black line. No relaxation: k-value equal to previous year estimates at the same period (black line). Prediction starts from July 1^st^, 2021 (vertical red line).

To focus on ICU hospitalizations, we estimated the ratio between ICU hospitalizations (from the SI-VIC database) and prevalent I_4_ cases in the past history of the pandemic (Figure S7), which is on average 3.62%. Whatever the scenario (**Figure 5**), the relaxation of barrier gestures leads to a predicted increase of prevalent ICU hospitalizations and the peak consistently exceeds French health care capacities (which is estimated at a maximum of about 5,000 ICU beds at the national level ^57^).

**Figure 5.**
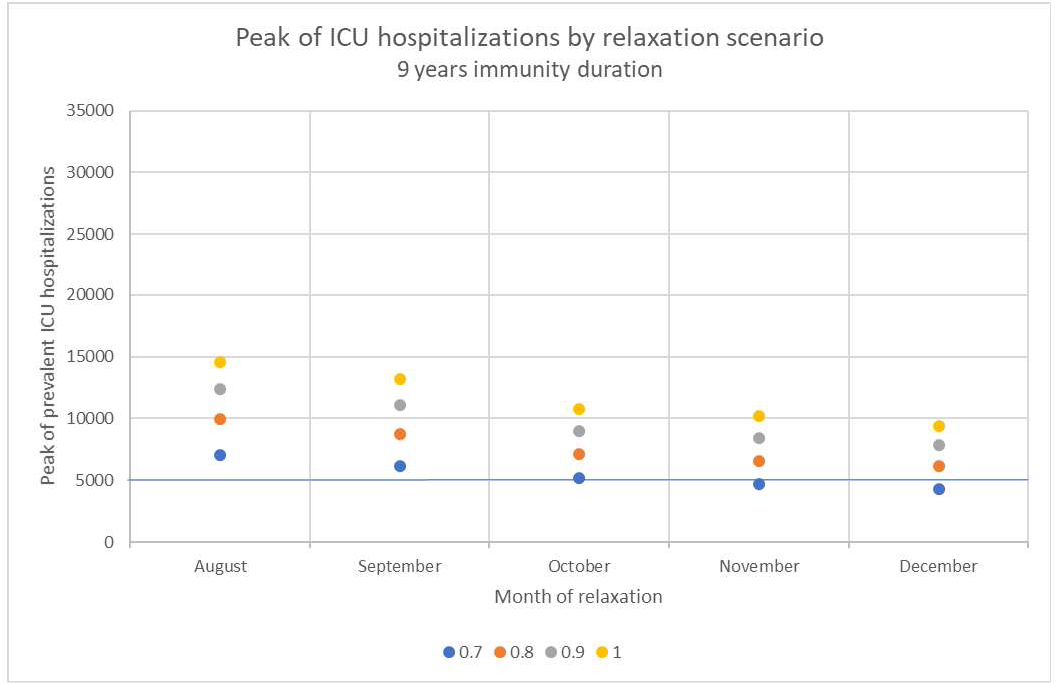
Peak of Intensive Care Units’ hospitalizations (ICU) predicted in France from July 2021 following each relaxation scenario, assuming that immunity wanes in 9 years. Relaxation scenarios: k-value (barrier gesture compliance index) raised either to 0.7, 0.8, 0.9 or 1 in August, September, October, November or December.

As expected, people over 75 years represent the major part of severe infections I_4_ (Figure S8). While 90% of the 75+ have been vaccinated at least once by the end of June 2021 (Figure S6), about 25% of them are fully susceptible in September 2021 (Figure S9) due both to waning immunity and to the vaccine efficacy hypothesis. Nevertheless, in a sensitivity analysis assuming a 90% vaccine efficacy against infections after 2 doses, even though the predicted peak ICU hospitalizations following each relaxation scenario is on average 41.0% lower than in the main analysis, a complete relaxation of barrier gestures still overwhelms health care system (Table S10 and Figure S10).

### 3.3. Impact of immunity duration hypothesis

As shown in **Figure 6**, our predictions are particularly sensitive to changes in our immunity duration assumptions. Assuming that immunity completely wanes over 3 years, we find that the ICU hospitalization peak is, on average, 86.1% higher than the same result when a 9-year immunity duration is assumed. In contrast, ICU hospitalizations peak at on average 46.1% lower when immunity is not allowed to wane (Table S11).

**Figure 6.**
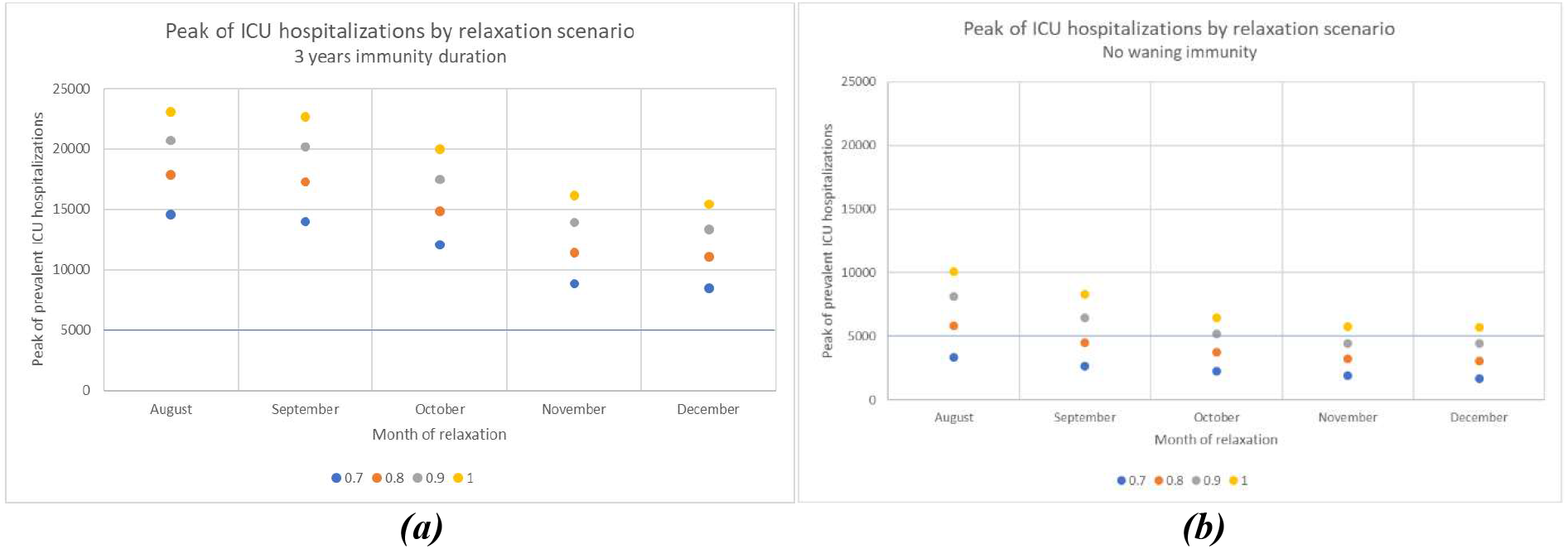
Peak of Intensive Care Units’ hospitalizations (ICU) predicted in France from July 2021 following each relaxation scenario, assuming that immunity wanes in 3 years **(a)**, or assuming that immunity does not wane **(b)**. Relaxation scenarios: k-value (barrier gesture compliance index) raised either to 0.7, 0.8, 0.9 or 1 in August, September, October, November or December.

## 3.4. Impact of vaccine hesitancy

In the absence of vaccine hesitancy, the vaccine coverage of at least one dose reaches 100% of the over 10-year-old population at the end of November 2021 (Figure S11 and Table S12). Under this assumption, ICU hospitalizations peak at, on average, 47.7% lower than in the main analysis, which includes vaccine hesitancy. This effect is particularly strong for the November and December relaxation scenarios, for which assuming no vaccine hesitancy reduces the hospitalization peak by about 65% (Table S13). In that case, a complete relaxation of barrier gestures would be possible in November and December 2021 without exceeding the French ICU hospitalization capacities (**Figure 7**).

**Figure 7.**
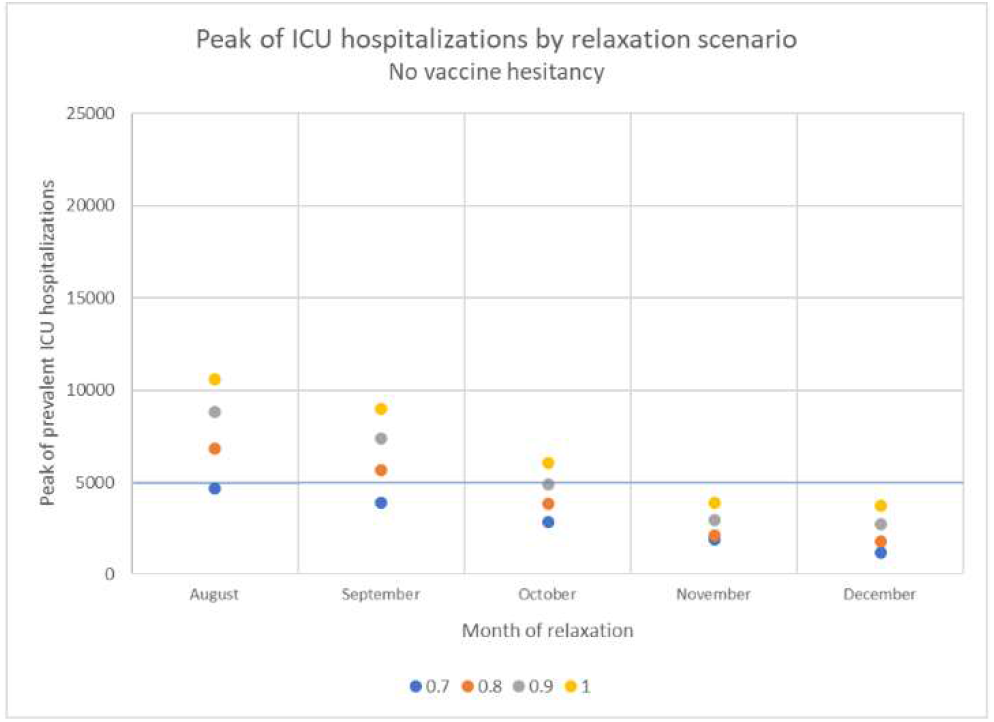
Peak of Intensive Care Units’ hospitalizations (ICU) predicted in France from July 2021 following each relaxation scenario assuming no vaccine hesitancy with a 9 years immunity duration. Relaxation scenarios: k-value (barrier gesture compliance index) raised either to 0.7, 0.8, 0.9 or 1 in August, September, October, November or December.

### 3.5. Booster campaign

Finally, we explored a crude implementation of a booster vaccination campaign for people over 75 years of age on the September 1^st^, 2021. In this analysis, we find a 16.3% average reduction in ICU peak hospitalization following barrier gesture relaxation (Table S14 and Figure S12). Even though this analysis is optimistic, in that it assumes that all but the vaccine hesitant people over 75+ will receive their booster dose on this date, this crude implementation of a booster dose allows us to estimate the best expected benefit from a booster vaccination campaign among the elderly.

## 4. Discussion

We used an age-structured compartmental model of COVID-19 that takes into account waning and boosting of immunity from vaccination and infection ^33^. We first showed that, considering the context of vaccine hesitancy in France and the delta variant breakthrough, maintaining application of barrier gestures appears essential to avoid a resurgence of severe infections that would exceed French health care capacities. Our study particularly highlighted the influence of immunity duration assumptions on the future short-term dynamic of the epidemic, which should not be neglected for further modelling. In addition, we reasserted the necessity to strengthen the vaccine rollout whereas current vaccines are effective (at least for severe disease) against the predominant delta variant ^58^. Indeed, among our sensitivity analyses, the only scenario that would allow a complete barrier gesture relaxation without overwhelming ICU beds capacities, is relaxation occurring from November with a 100% vaccine coverage of the eligible population. Finally, according to our very preliminary analysis, the strategy consisting in the revaccination of people over 75, would not allow us to relax barrier gestures safely, in particular in the context of vaccine hesitancy. Nevertheless, further modelling is needed to evaluate more accurately this last statement.

Our work is consistent with other French modelling having estimated that, given the current vaccine rollout and the progression of the delta variant, barrier gesture application will be necessary to maintain severe cases under health care capacities’ limits ^59 60^. Nevertheless, our predictions tend to be more pessimistic. One explication is that we took into account the greater transmissibility of the delta variant at the level of that it has been estimated by ECDC ^29^, whereas these studies explored an inferior range of effective reproductive numbers. In addition, these studies assumed that immunity acquired after an infection or vaccination is definitive. Finally, our model, incorporating contact patterns, takes into account both school re-opening and recovery of on-site work, which leads to an increased transmission rates in September. Similar to our findings, a very high vaccination coverage has been estimated to be required to allow a complete relaxation of control measures ^28^ and a high vaccine hesitancy has been related to a prolonged need for non-pharmaceutical-interventions ^61^. On the other hand, our work highlighted the influence of the immunity duration on the short-term dynamic of the epidemic. Few other studies chose to modelling the COVID-19 epidemic incorporating waning immunity ^23 24 25 26 33^. Giannitsarou et al. estimated that, given the population turnover, the disease would become endemic in the long term with recurrent waves, which magnitude would depends on how fast immunity wanes ^26^. Saad-Roy et al. showed that, the shorter immunity duration conferred by vaccination, the higher the vaccine coverage required to achieve herd immunity ^25^.

Measuring compliance of the population to barrier gestures is challenging. Our model is fit using a single parameter, the k-value. Because we incorporated the changes in contact patterns consequently to governmental measures directly in the contact matrices, this k-value reflects the population compliance to barrier gestures. Nevertheless, it also incorporates temporal variations in testing and variations in climatic conditions which influence transmission ^8^. However, even though the detection rate increased significantly during the first months of the epidemic, it remained relatively stable during our calibration period ^62^. Concerning climatic conditions, we estimated in another modelling study that transmission is increased in average by 10% during the winter period and decreased by 22% during the summer period ^8^. Because we did not account for these variations, we expect our predictions to be somewhat overestimated in summer and underestimated in winter. Another limitation is that our model underestimates vaccine efficacy against severe infections after 2 doses, assuming that it is equal to vaccine efficacy against all infections (which is set to 80% in the main analysis). However, assuming an optimistic 90% vaccine efficacy after 2 doses against infections and severe cases, leads to similar conclusions with no possibility to fully relax barrier gestures safely. Moreover, we did not account for the increased risk of hospitalization related to the alpha variant compared to the historical strain ^63^ which could explain that our ICU hospitalizations/prevalent I_4_ ratio tends to increase over the calibration period. This could lead to an underestimation of the ICU hospitalizations rebound. Finally, we did not account for the age-related immune response heterogeneity to SARS-CoV2 infection and vaccine, which will have to be considered in the future to investigate more accurately the impact of a booster vaccination campaign among elderly.

In conclusion, while the durability of immunity provided by an infection or by vaccination remains uncertain, we provide an insight on the predictable dynamic of the epidemic in France assuming that immunity wanes. To date, maintaining the application of barrier gestures seems unavoidable in the French context to not overwhelm health care capacities. While the possibility of a booster campaign among elderly is being considered, scientific evidences on its potential benefit are lacking.

## Data Availability

Variants of concern data, reported COVID-19 cases data and prevalent intensive care units hospitalization data for France are publicly available online.

https://www.data.gouv.fr/fr/datasets/donnees-de-laboratoires-pour-le-depistage-indicateurs-sur-les-variants/

https://www.data.gouv.fr/fr/datasets/donnees-relatives-aux-resultats-des-tests-virologiques-covid-19/

https://geodes.santepubliquefrance.fr/#c=indicator&f=0&i=covid_hospit.rea&s=2021-08-03&t=a01&view=map2

## Author Contributions

Methodology, J.H., M.P., L.W. R.T.; software, D.D.; formal analysis, C.V.; writing— original draft preparation, C.V.; writing—review and editing, C.V., D.D., J.H., L.W., M.P., R.T.; All authors have read and agreed to the published version of the manuscript.

## Acknowledgements

We thank Lauren Childs, Jing Li, Zhilan Feng, John Glasser and Gergely Rost for their discussions and input on this project.

## Funding

This research received no external funding.

## Data Availability Statement

Variants of concern data for France are publicly available on: *https://www.data.gouv.fr/fr/datasets/donnees-de-laboratoires-pour-le-depistage-indicateurs-sur-les-variants/*

Reported COVID-19 cases data for France are available on: *https://www.data.gouv.fr/fr/datasets/donnees-relatives-aux-resultats-des-tests-virologiques-covid-19/*

Prevalent intensive care unit’s hospitalization data are for France are publicly available on: *https://geodes.santepubliquefrance.fr/#c=indicator&f=0&i=covid_hospit.rea&s=2021-08-03&t=a01&view=map2*

## Conflicts of Interest

The authors declare no conflict of interest.

## Appendix A

## Model equations

Our model tracks age, infection and immune status. Susceptible individuals of status i and age m are denoted by S_im_. Infectious individuals of symptom severity j and age m are denoted by I_jm_. Infected but not-yet-infectious individuals of symptom severity j, age m and stage k are denoted by E^k^_jm_. Vaccinated individuals of initial immune status i, age m and dose l are denoted by V^l^_im_. Parameter descriptions are found in **Table 1** in the main test. The system of ODEs for age group m is given by the following set of equations:

### Susceptible compartments

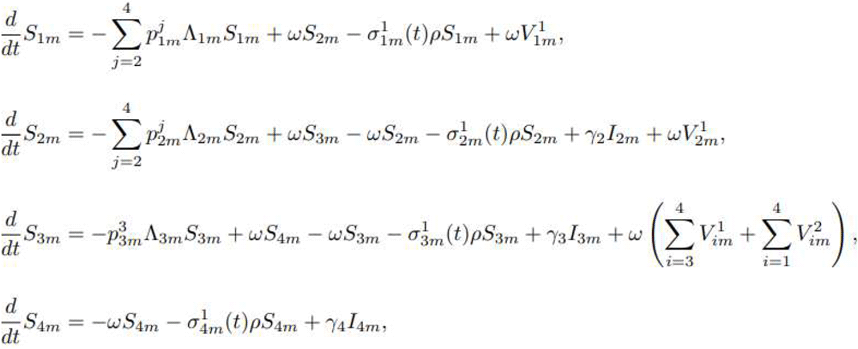

### Vaccinated compartments

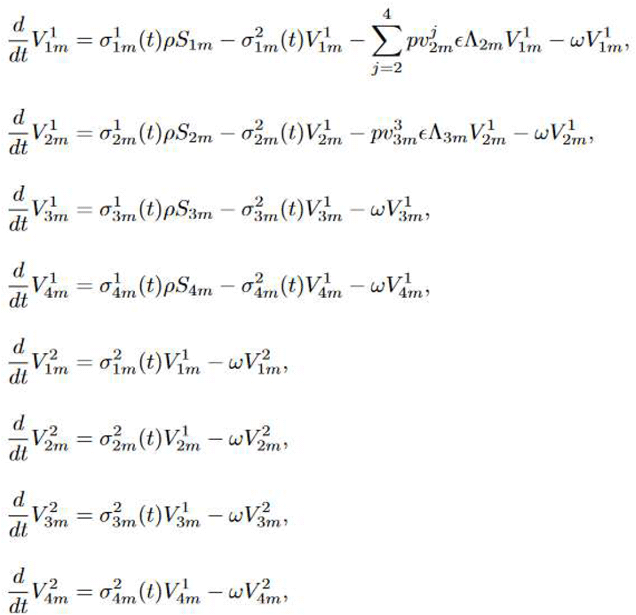

### Exposed compartment

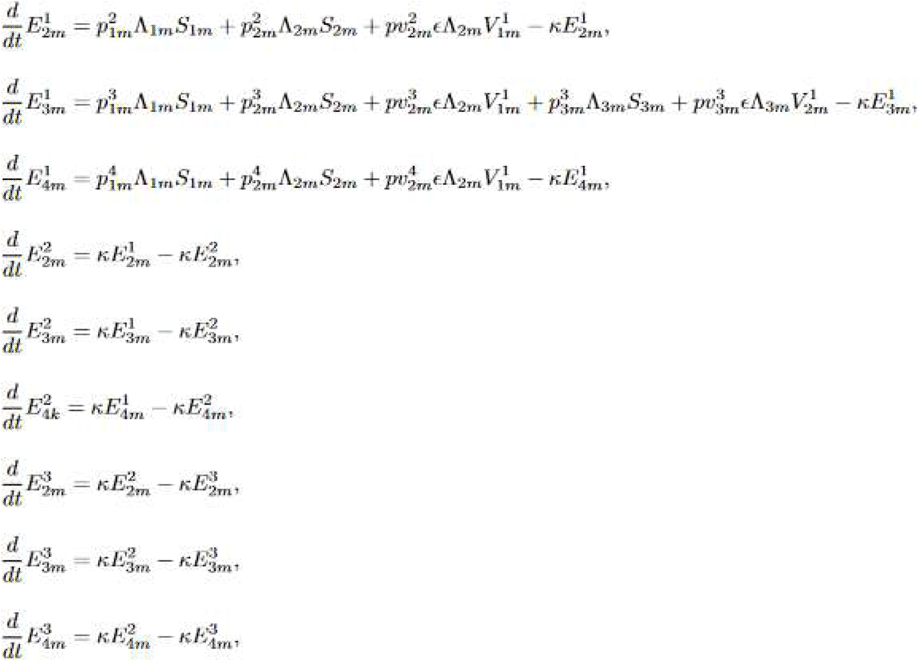

Where,

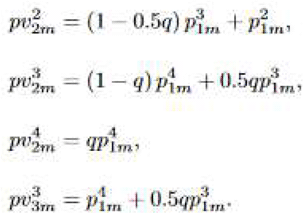

### Infectious compartment

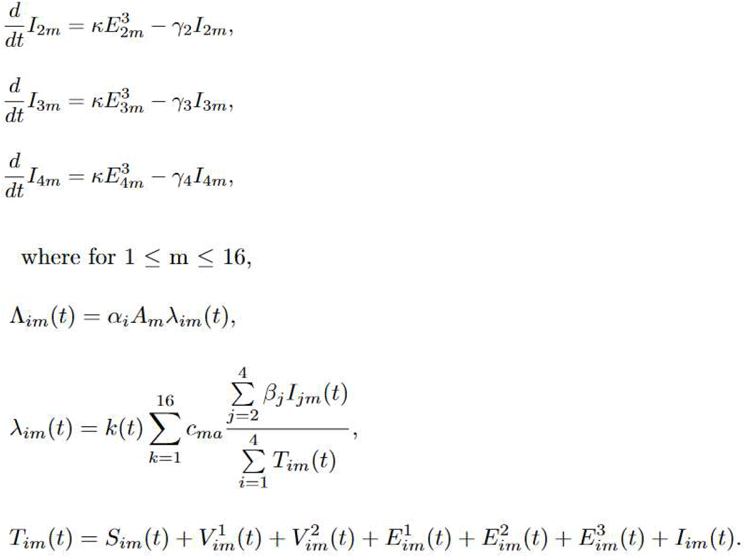

## Notes

### Competing Interest Statement

The authors have declared no competing interest.

### Author Declarations

A formal ethics approval was not needed for our modelling study.

