## Supplementary materials for "Barrier gesture relaxation during vaccination campaign in France: modelling impact of waning immunity"

***Table S1.*** *Size of the population in each age group from INSEE French census (Institut National de la Statistique et des Etudes Economiques) as of January 1st, 2021*

| Age class | Size |
|---|---|
| [0;4] | 3,671,719 |
| [5;9] | 4,084,036 |
| [10;14] | 4,187,992 |
| [15;19] | 4,140,996 |
| [20;24] | 3,757,482 |
| [25;29] | 3,713,426 |
| [30;34] | 4,056,469 |
| [35;39] | 4,231,788 |
| [40;44] | 4,072,226 |
| [45;49] | 4,512,223 |
| [50;54] | 4,425,730 |
| [55;59] | 4,359,376 |
| [60;64] | 4,099,662 |
| [65;69] | 3,899,944 |
| [70;74] | 3,477,098 |
| ≥ 75 | 6,373,536 |
| Total | 67,063,703 |

***Table S2****. Proportions of mild ($p^2_{im}$), moderate ($p^3_{im}$) and severe ($p^4_{im}$) infections by age m ($1 \leq m \leq 16$) and by susceptibility status i ($1 \leq i \leq 4$)*

| Age class (m) | $p^2_{1m}$ | $p^3_{1m}$ | $p^4_{1m}$ |
|---|---|---|---|
| [0;4] | 0,985 | 0,014 | 0,001 |
| [5;9] | 0,979 | 0,020 | 0,001 |
| [10;14] | 0,976 | 0,023 | 0,001 |
| [15;19] | 0,960 | 0,038 | 0,003 |
| [20;24] | 0,930 | 0,065 | 0,005 |
| [25;29] | 0,900 | 0,091 | 0,009 |
| [30;34] | 0,874 | 0,112 | 0,014 |
| [35;39] | 0,848 | 0,132 | 0,020 |
| [40;44] | 0,808 | 0,162 | 0,030 |
| [45;49] | 0,766 | 0,191 | 0,043 |
| [50;54] | 0,710 | 0,227 | 0,064 |
| [55;59] | 0,622 | 0,281 | 0,098 |
| [60;64] | 0,527 | 0,331 | 0,143 |
| [65;69] | 0,435 | 0,367 | 0,197 |
| [70;74] | 0,348 | 0,390 | 0,262 |
| ≥ 75 | 0,218 | 0,364 | 0,418 |

$p^j_{1m}$ are directly informed by [35].

| Age class (m) | $p^2_{2m}$ | $p^3_{2m}$ | $p^4_{2m}$ |
|---|---|---|---|
| [0;4] | 0,9969 | 0,0028 | 0,0003 |
| [5;9] | 0,9960 | 0,0037 | 0,0003 |
| [10;14] | 0,9956 | 0,0042 | 0,0003 |
| [15;19] | 0,9923 | 0,0078 | 0,0009 |
| [20;24] | 0,9853 | 0,0133 | 0,0015 |
| [25;29] | 0,9774 | 0,0200 | 0,0027 |
| [30;34] | 0,9692 | 0,0266 | 0,0042 |
| [35;39] | 0,9602 | 0,0338 | 0,0060 |
| [40;44] | 0,9457 | 0,0453 | 0,0090 |
| [45;49] | 0,9284 | 0,0588 | 0,0129 |
| [50;54] | 0,9030 | 0,0789 | 0,0192 |
| [55;59] | 0,8609 | 0,1108 | 0,0294 |
| [60;64] | 0,8084 | 0,1498 | 0,0429 |
| [65;69] | 0,7470 | 0,1930 | 0,0591 |
| [70;74] | 0,6795 | 0,2419 | 0,0786 |
| ≥ 75 | 0,5274 | 0,3472 | 0,1254 |

$p^j_{2m}$ are calculated assuming that $p^2_{2m} = p^2_{1m} + (1-0.5*0.3)p^3_{1m}$ , $p^3_{2m} = 0.7*p^4_{1m} + 0.5*0.3*p^3_{1m}$ and $p^4_{2m} = 0.3*p^4_{1m}$.

| Age class (m) | $p^2_{3m}$ | $p^3_{3m}$ | $p^4_{3m}$ |
|---|---|---|---|
| [0;4] | 0,985 | 0,015 | 0,000 |
| [5;9] | 0,979 | 0,021 | 0,000 |
| [10;14] | 0,976 | 0,024 | 0,000 |
| [15;19] | 0,960 | 0,041 | 0,000 |
| [20;24] | 0,930 | 0,070 | 0,000 |
| [25;29] | 0,900 | 0,100 | 0,000 |
| [30;34] | 0,874 | 0,126 | 0,000 |
| [35;39] | 0,848 | 0,152 | 0,000 |
| [40;44] | 0,808 | 0,192 | 0,000 |
| [45;49] | 0,766 | 0,234 | 0,000 |
| [50;54] | 0,710 | 0,291 | 0,000 |
| [55;59] | 0,622 | 0,379 | 0,000 |
| [60;64] | 0,527 | 0,474 | 0,000 |
| [65;69] | 0,435 | 0,564 | 0,000 |
| [70;74] | 0,348 | 0,652 | 0,000 |
| ≥ 75 | 0,218 | 0,782 | 0,000 |

$p^j_{3m}$ are calculated assuming that $p^2_{3m} = p^2_{1m}$, $p^3_{3m} = p^4_{1m} + p^3_{1m}$ and $p^4_{3m} = 0$.

***Table S3.*** *Eligible age classes to vaccination by month*

| Month | Eligible age class | Month | Eligible age class |
|---|---|---|---|
| January | ≥ 75 | July | ≥ 10 |
| February | ≥ 75 | August | ≥ 10 |
| March | ≥ 60 | September | ≥ 10 |
| April | ≥ 50 | October | ≥ 10 |
| May | ≥ 20 | November | ≥ 10 |
| June | ≥ 10 | December | ≥ 10 |

***Table S4.*** *Vaccine coverage with at least one dose at the end of each month informed by vaccination French database VAC-SI (Système d'Informations pour le suivi de la VACcination)*

| Month | Vaccine coverage with at least one dose at the end of each month (%) |
|---|---|
| January | 2.45 |
| February | 4.60 |
| March | 12.22 |
| April | 23.63 |
| May | 38.85 |
| June | 50.64 |

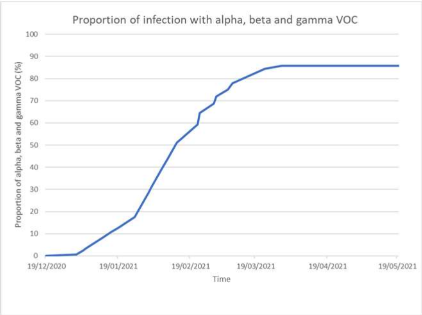


***Figure S1.*** *Proportion of infections by alpha, beta or gamma Variants Of Concern (VOC) from French surveillance database SI-DEP (Système d'Informations de DEPistage) and Santé Publique France surveys from January 2021 to May 2021*

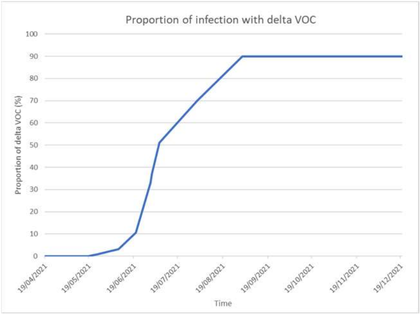


***Figure S2.*** *Proportion of infections by delta Variant Of Concern (VOC) from French surveillance database SI-DEP (Système d'Informations de DEPistage) from May 2021 to July 2021 and from ECDC's (European Centre for Disease Prevention and Control) projections from July to August 2021*

***Table S5.*** *Contact matrices for pre-pandemic contacts at home, at school, at work and other contacts for France, provided by Prem et. al* [40]

**HOME**

| | 0 | 5 | 10 | 15 | 20 | 25 | 30 | 35 | 40 | 45 | 50 | 55 | 60 | 65 | 70 | 75 |
|---|---|---|---|---|---|---|---|---|---|---|---|---|---|---|---|---|
| **0** | 0,688061 | 0,47768 | 0,19942 | 0,080462 | 0,134592 | 0,379075 | 0,702365 | 0,571827 | 0,198565 | 0,066211 | 0,052676 | 0,036809 | 0,02239 | 0,006494 | 0,002993 | 0,00522 |
| **5** | 0,325932 | 1,009634 | 0,385944 | 0,126838 | 0,037585 | 0,145181 | 0,573297 | 0,714298 | 0,468638 | 0,129078 | 0,03717 | 0,022007 | 0,011999 | 0,006392 | 0,002603 | 0,003312 |
| **10** | 0,128181 | 0,369161 | 1,588507 | 0,421811 | 0,059928 | 0,026051 | 0,126748 | 0,457233 | 0,63762 | 0,272502 | 0,063979 | 0,014102 | 0,008759 | 0,0083 | 0,006661 | 0,003335 |
| **15** | 0,047173 | 0,096142 | 0,392455 | 1,254115 | 0,206093 | 0,032457 | 0,017091 | 0,168149 | 0,401699 | 0,495991 | 0,189244 | 0,042055 | 0,009506 | 0,007754 | 0,00499 | 0,002221 |
| **20** | 0,096405 | 0,047508 | 0,072079 | 0,390331 | 1,191402 | 0,197242 | 0,04027 | 0,01859 | 0,117882 | 0,487593 | 0,271308 | 0,112326 | 0,014587 | 0,002322 | 0,0024 | 0,002529 |
| **25** | 0,366726 | 0,096787 | 0,025564 | 0,055108 | 0,207189 | 1,014666 | 0,195743 | 0,028819 | 0,013956 | 0,05159 | 0,209117 | 0,098403 | 0,037097 | 0,007793 | 0,000856 | 0,004342 |
| **30** | 0,519928 | 0,475619 | 0,163414 | 0,023519 | 0,046657 | 0,19064 | 0,932506 | 0,199322 | 0,053371 | 0,015524 | 0,030213 | 0,051241 | 0,050143 | 0,007601 | 0,003313 | 0,003024 |
| **35** | 0,447115 | 0,711948 | 0,552992 | 0,193906 | 0,023157 | 0,030994 | 0,163766 | 1,042591 | 0,164947 | 0,03814 | 0,019008 | 0,014456 | 0,027545 | 0,012462 | 0,006892 | 0,002375 |
| **40** | 0,169285 | 0,41168 | 0,661424 | 0,441584 | 0,088938 | 0,022194 | 0,059488 | 0,159125 | 0,867285 | 0,144631 | 0,028336 | 0,004299 | 0,015838 | 0,014718 | 0,005351 | 0,005975 |
| **45** | 0,08251 | 0,180596 | 0,361976 | 0,618391 | 0,36123 | 0,051807 | 0,02151 | 0,064652 | 0,144427 | 0,796167 | 0,124662 | 0,0233 | 0,007601 | 0,00722 | 0,005596 | 0,008886 |
| **50** | 0,125564 | 0,103706 | 0,201282 | 0,338546 | 0,390068 | 0,230417 | 0,065329 | 0,033642 | 0,060287 | 0,166583 | 0,74747 | 0,132954 | 0,021398 | 0,00577 | 0,004745 | 0,021755 |
| **55** | 0,233811 | 0,217834 | 0,146832 | 0,233026 | 0,270983 | 0,290321 | 0,213433 | 0,060539 | 0,027171 | 0,107779 | 0,221376 | 0,866688 | 0,125525 | 0,034265 | 0,004821 | 0,01819 |
| **60** | 0,261257 | 0,23384 | 0,163821 | 0,135336 | 0,107688 | 0,169806 | 0,261427 | 0,158204 | 0,075378 | 0,033413 | 0,086644 | 0,193533 | 0,908575 | 0,109923 | 0,022561 | 0,0055 |
| **65** | 0,139879 | 0,214595 | 0,200491 | 0,107827 | 0,065775 | 0,082189 | 0,144265 | 0,149013 | 0,127643 | 0,055598 | 0,047119 | 0,079539 | 0,126782 | 0,691952 | 0,090216 | 0,011954 |
| **70** | 0,081573 | 0,242322 | 0,234412 | 0,213315 | 0,031554 | 0,065307 | 0,060626 | 0,164654 | 0,141832 | 0,130553 | 0,062974 | 0,036982 | 0,115141 | 0,185317 | 0,52601 | 0,108659 |
| **75** | 0,199006 | 0,248851 | 0,384293 | 0,301761 | 0,073838 | 0,074449 | 0,094919 | 0,190461 | 0,264034 | 0,204618 | 0,357401 | 0,140654 | 0,055149 | 0,103625 | 0,110188 | 0,42997 |

**SCHOOL**

| | 0 | 5 | 10 | 15 | 20 | 25 | 30 | 35 | 40 | 45 | 50 | 55 | 60 | 65 | 70 | 75 |
|---|---|---|---|---|---|---|---|---|---|---|---|---|---|---|---|---|
| **0** | 2,423317 | 0,316795 | 0,056616 | 0,061873 | 0,016538 | 0,086418 | 0,159975 | 0,13168 | 0,062385 | 0,073877 | 0,039413 | 0,026864 | 0,002379 | 0,000823 | 8,27E-66 | 6,40E-120 |
| **5** | 0,396403 | 2,693404 | 0,149118 | 0,016606 | 0,01669 | 0,055242 | 0,074119 | 0,07873 | 0,07744 | 0,055006 | 0,04699 | 0,015159 | 0,004197 | 0,001096 | 0,000348 | 8,10E-39 |
| **10** | 0,003278 | 0,57892 | 3,343917 | 0,10431 | 0,01068 | 0,042896 | 0,044908 | 0,080341 | 0,085068 | 0,066506 | 0,047289 | 0,024727 | 0,00449 | 0,000572 | 4,94E-25 | 0,000182 |
| **15** | 0,019842 | 0,024969 | 1,045318 | 3,605976 | 0,043691 | 0,048554 | 0,055761 | 0,08817 | 0,075296 | 0,088394 | 0,049428 | 0,030491 | 0,004869 | 0,000825 | 6,22E-33 | 1,71E-70 |
| **20** | 0,022229 | 0,013345 | 0,005293 | 0,446844 | 0,257036 | 0,033255 | 0,022805 | 0,030292 | 0,019724 | 0,024325 | 0,013184 | 0,00966 | 0,000622 | 0,001128 | 0,000177 | 1,22E-47 |
| **25** | 0,027886 | 0,076988 | 0,02407 | 0,124165 | 0,184199 | 0,126952 | 0,022679 | 0,032915 | 0,040019 | 0,035284 | 0,009371 | 0,015596 | 0,004081 | 0,002166 | 0,000463 | 0,001287 |
| **30** | 0,056356 | 0,264675 | 0,174398 | 0,110956 | 0,03436 | 0,070986 | 0,065925 | 0,050138 | 0,0559 | 0,029315 | 0,024374 | 0,003783 | 0,005294 | 0,000404 | 1,66E-48 | 3,11E-55 |
| **35** | 0,105979 | 0,17561 | 0,128075 | 0,065077 | 0,014619 | 0,050071 | 0,071387 | 0,058714 | 0,067007 | 0,032327 | 0,004871 | 0,011483 | 0,000584 | 0,001916 | 1,85E-123 | 9,71E-67 |
| **40** | 0,034648 | 0,102567 | 0,081044 | 0,326451 | 0,007181 | 0,025663 | 0,025759 | 0,038373 | 0,084545 | 0,030394 | 0,034463 | 0,00974 | 0,005925 | 0,000443 | 4,82E-68 | 2,41E-92 |
| **45** | 0,290592 | 0,230712 | 0,148747 | 0,530154 | 0,005225 | 0,032159 | 0,063435 | 0,060411 | 0,057917 | 0,035105 | 0,043871 | 0,020366 | 0,003608 | 0,001577 | 6,22E-134 | 3,28E-72 |
| **50** | 0,071845 | 0,385147 | 0,479616 | 0,511824 | 0,00535 | 0,016632 | 0,047436 | 0,051439 | 0,065808 | 0,090621 | 0,046598 | 0,025021 | 0,005172 | 8,47E-24 | 1,24E-117 | 5,65E-78 |
| **55** | 0,222714 | 0,373913 | 0,322418 | 0,365212 | 0,006354 | 0,065086 | 0,025817 | 0,047263 | 0,061043 | 0,044548 | 0,039369 | 0,043929 | 0,010233 | 1,10E-31 | 0,000783 | 0,000763 |
| **60** | 0,067492 | 0,052495 | 0,026763 | 0,124012 | 0,011289 | 0,001616 | 0,015128 | 0,044256 | 0,009915 | 0,015818 | 0,012666 | 0,006649 | 0,020707 | 0,010852 | 4,42E-67 | 2,12E-37 |
| **65** | 0,001774 | 0,021404 | 0,008328 | 5,82E-32 | 0,00177 | 0,001728 | 0,011233 | 0,004955 | 0,005022 | 0,008109 | 0,001858 | 0,014481 | 0,007931 | 0,017278 | 0,011123 | 3,46E-126 |
| **70** | 1,29E-28 | 5,11E-26 | 1,93E-40 | 0,007609 | 2,64E-22 | 1,70E-24 | 1,26E-26 | 0,007628 | 0,007858 | 0,021177 | 0,035255 | 0,02146 | 0,007741 | 0,008013 | 0,007913 | 0,021383 |
| **75** | 2,83E-94 | 0,021161 | 8,48E-42 | 0,021289 | 4,90E-36 | 0,007598 | 9,78E-69 | 2,23E-60 | 1,44E-48 | 8,57E-60 | 4,70E-42 | 1,60E-46 | 2,21E-83 | 8,86E-107 | 1,02E-80 | 6,61E-113 |

**WORK**

| | 0 | 5 | 10 | 15 | 20 | 25 | 30 | 35 | 40 | 45 | 50 | 55 | 60 | 65 | 70 | 75 |
|---|---|---|---|---|---|---|---|---|---|---|---|---|---|---|---|---|
| **0** | 0 | 0 | 0 | 0 | 0 | 0 | 0 | 0 | 0 | 0 | 0 | 0 | 0 | 8,21E-92 | 1,21E-05 | 3,16E-125 |
| **5** | 0 | 0 | 0 | 0 | 0 | 0 | 0 | 0 | 0 | 0 | 0 | 0 | 0 | 1,35E-05 | 7,65E-79 | 2,38E-65 |
| **10** | 0 | 0 | 0,014566 | 0,006934 | 0,010019 | 0,002899 | 0,02392 | 0,006819 | 0,024009 | 0,013956 | 0,005312 | 5,66E-09 | 1,58E-18 | 2,80E-53 | 4,96E-06 | 3,78E-102 |
| **15** | 0 | 0 | 0,012359 | 0,465661 | 0,504972 | 0,290428 | 0,273328 | 0,254158 | 0,284017 | 0,224052 | 0,126467 | 0,025944 | 0,001402 | 8,35E-06 | 2,86E-06 | 1,89E-31 |
| **20** | 0 | 0 | 0,021889 | 0,340347 | 0,891532 | 0,854474 | 0,719952 | 0,793497 | 0,60096 | 0,481606 | 0,353457 | 0,070214 | 0,005224 | 9,86E-06 | 1,33E-05 | 3,74E-06 |
| **25** | 0 | 0 | 0,028152 | 0,29917 | 0,848489 | 1,428921 | 1,032235 | 1,008132 | 0,934231 | 0,655398 | 0,517834 | 0,103426 | 0,006969 | 1,61E-05 | 1,01E-05 | 3,01E-06 |
| **30** | 0 | 0 | 0,032201 | 0,163215 | 0,593526 | 0,978649 | 1,292918 | 1,134236 | 1,019614 | 0,832958 | 0,482937 | 0,121815 | 0,006588 | 1,64E-05 | 4,10E-06 | 3,49E-06 |
| **35** | 0 | 0 | 0,019809 | 0,325651 | 0,492504 | 0,9195 | 0,994002 | 1,373982 | 1,340825 | 0,928444 | 0,649787 | 0,111775 | 0,00467 | 1,23E-05 | 9,14E-06 | 6,02E-06 |
| **40** | 0 | 0 | 0,022453 | 0,204014 | 0,581487 | 0,912221 | 1,086205 | 1,162794 | 1,428988 | 1,148896 | 0,793711 | 0,119043 | 0,006869 | 1,44E-05 | 1,03E-05 | 1,30E-05 |
| **45** | 0 | 0 | 0,029976 | 0,25517 | 0,396557 | 0,686163 | 0,888148 | 0,976758 | 1,013636 | 0,989896 | 0,612885 | 0,13482 | 0,005475 | 1,63E-05 | 1,08E-05 | 6,09E-06 |
| **50** | 0 | 0 | 0,03057 | 0,180021 | 0,311334 | 0,652049 | 0,767315 | 0,799289 | 1,085675 | 1,047858 | 0,765183 | 0,160481 | 0,006171 | 1,18E-05 | 1,18E-05 | 1,02E-05 |
| **55** | 0 | 0 | 0,020975 | 0,053869 | 0,093412 | 0,171829 | 0,241186 | 0,227484 | 0,291715 | 0,230261 | 0,205195 | 0,055507 | 0,002592 | 1,35E-05 | 6,59E-06 | 6,66E-06 |
| **60** | 0 | 0 | 0,001689 | 0,001742 | 0,008855 | 0,015784 | 0,017553 | 0,020751 | 0,022507 | 0,021991 | 0,01615 | 0,005389 | 0,000212 | 2,03E-05 | 8,26E-06 | 1,48E-05 |
| **65** | 7,60E-06 | 3,36E-06 | 7,65E-06 | 2,28E-05 | 3,15E-05 | 7,89E-05 | 7,24E-05 | 2,92E-05 | 6,62E-05 | 5,96E-05 | 7,71E-05 | 5,31E-05 | 4,66E-05 | 1,42E-05 | 2,49E-05 | 1,19E-05 |
| **70** | 5,79E-55 | 7,89E-42 | 2,55E-06 | 2,61E-05 | 1,68E-05 | 2,12E-05 | 3,57E-05 | 4,02E-05 | 3,56E-05 | 3,10E-05 | 2,13E-05 | 4,50E-05 | 2,61E-05 | 1,68E-05 | 1,67E-05 | 2,61E-05 |
| **75** | 2,36E-141 | 9,07E-97 | 1,19E-89 | 9,40E-22 | 4,66E-05 | 4,70E-05 | 4,69E-05 | 8,42E-05 | 2,78E-05 | 1,03E-05 | 1,07E-05 | 7,26E-75 | 1,10E-65 | 1,03E-05 | 5,17E-49 | 8,28E-43 |

**OTHER**

| | 0 | 5 | 10 | 15 | 20 | 25 | 30 | 35 | 40 | 45 | 50 | 55 | 60 | 65 | 70 | 75 |
|---|---|---|---|---|---|---|---|---|---|---|---|---|---|---|---|---|
| **0** | 0,693543 | 0,311971 | 0,155879 | 0,113034 | 0,190623 | 0,287716 | 0,346709 | 0,338489 | 0,234633 | 0,170371 | 0,214966 | 0,208538 | 0,161568 | 0,120728 | 0,084797 | 0,048645 |
| **5** | 0,339689 | 1,329525 | 0,475759 | 0,12955 | 0,109453 | 0,214681 | 0,2733 | 0,345402 | 0,299103 | 0,14732 | 0,123148 | 0,148272 | 0,160718 | 0,104148 | 0,055309 | 0,049799 |
| **10** | 0,106863 | 0,590358 | 2,038973 | 0,312856 | 0,206428 | 0,172536 | 0,207677 | 0,282532 | 0,333989 | 0,217512 | 0,149173 | 0,107785 | 0,08828 | 0,079298 | 0,063044 | 0,066149 |
| **15** | 0,057277 | 0,187355 | 0,851206 | 2,505921 | 0,605197 | 0,279758 | 0,184743 | 0,251656 | 0,263827 | 0,240537 | 0,113363 | 0,065412 | 0,055955 | 0,044238 | 0,029544 | 0,023918 |
| **20** | 0,081116 | 0,107359 | 0,156286 | 0,987959 | 1,593821 | 0,62267 | 0,384748 | 0,290762 | 0,231479 | 0,276742 | 0,175452 | 0,135016 | 0,063394 | 0,042305 | 0,051882 | 0,04984 |
| **25** | 0,138575 | 0,074855 | 0,065801 | 0,252517 | 0,70375 | 0,88757 | 0,526248 | 0,400917 | 0,274421 | 0,254707 | 0,224766 | 0,140037 | 0,064387 | 0,041678 | 0,029247 | 0,017978 |
| **30** | 0,145962 | 0,107803 | 0,159891 | 0,125116 | 0,330466 | 0,480173 | 0,62795 | 0,510084 | 0,328597 | 0,257376 | 0,272532 | 0,228553 | 0,122083 | 0,077394 | 0,043692 | 0,049512 |
| **35** | 0,150701 | 0,184586 | 0,135759 | 0,098644 | 0,234417 | 0,406634 | 0,530645 | 0,735539 | 0,525019 | 0,345444 | 0,245835 | 0,226396 | 0,226892 | 0,13801 | 0,091157 | 0,043993 |
| **40** | 0,105601 | 0,142397 | 0,217832 | 0,137317 | 0,245897 | 0,291815 | 0,419986 | 0,516326 | 0,573295 | 0,357911 | 0,25638 | 0,140133 | 0,154903 | 0,095672 | 0,081612 | 0,042024 |
| **45** | 0,032707 | 0,055871 | 0,071064 | 0,133008 | 0,182393 | 0,22377 | 0,286147 | 0,363672 | 0,357179 | 0,375306 | 0,304886 | 0,162199 | 0,134615 | 0,077554 | 0,077503 | 0,072987 |
| **50** | 0,054309 | 0,105924 | 0,117409 | 0,171282 | 0,312061 | 0,391509 | 0,299175 | 0,330171 | 0,382468 | 0,438002 | 0,359199 | 0,33071 | 0,234526 | 0,112724 | 0,083306 | 0,068495 |
| **55** | 0,085911 | 0,079442 | 0,083446 | 0,097191 | 0,242813 | 0,404515 | 0,437103 | 0,412624 | 0,41284 | 0,298282 | 0,456976 | 0,495798 | 0,374599 | 0,183981 | 0,114336 | 0,075718 |
| **60** | 0,067381 | 0,076573 | 0,064345 | 0,100325 | 0,210947 | 0,320841 | 0,345277 | 0,470832 | 0,41564 | 0,345055 | 0,325992 | 0,46949 | 0,476478 | 0,305696 | 0,240407 | 0,123039 |
| **65** | 0,056358 | 0,077436 | 0,048044 | 0,049635 | 0,144551 | 0,223139 | 0,294537 | 0,300319 | 0,29264 | 0,238548 | 0,275224 | 0,346345 | 0,383277 | 0,293799 | 0,174542 | 0,127017 |
| **70** | 0,023676 | 0,052167 | 0,073379 | 0,134391 | 0,124321 | 0,179175 | 0,192428 | 0,304729 | 0,365257 | 0,282239 | 0,240968 | 0,279666 | 0,537069 | 0,42586 | 0,440137 | 0,195697 |
| **75** | 0,044552 | 0,047109 | 0,067932 | 0,036831 | 0,080415 | 0,106128 | 0,210116 | 0,191881 | 0,187383 | 0,238243 | 0,181391 | 0,203428 | 0,224651 | 0,263519 | 0,257358 | 0,212948 |

***Table S6.*** *Definitions of perturbation matrices for contacts at school (S), at work (W) and other contacts (O)*

| Perturbation matrices | Definitions |
|---|---|
| S2 | School closure or national vacations |
| S3 | Zone A vacations |
| S4 | Zones A and C vacations |
| S5 | Zones B and C vacations |
| S6 | Zone B vacations |
| O1 | Closure of bars/restaurants, cinemas/theatres, non-essential businesses and travels restriction < 3 hours to < 20km from home (2nd and 3rd lockdowns) |
| O2 | Closure of bars/restaurants, cinemas/theatres, non-essential businesses and strict travels restriction < 1 hours à < 1km (1st lockdown) |
| O3 | Closure of bars/restaurants, cinemas/theatres and curfew |
| W1 | Obligatory to work from home if feasible (2nd and 3rd lockdowns) |
| W2 | Strict obligatory to work from home, lay-off if not feasible (1st lockdown) |
| W3 | Recommendation to work from home |

*Zone A: academies of Besançon, Bordeaux, Clermont-Ferrand, Dijon, Grenoble, Limoges, Lyon and Poitiers/ Zone B : academies of Aix-Marseille, Amiens, Caen, Lille, Nancy-Metz, Nantes, Nice, Orléans-Tours, Reims, Rennes, Rouen and Strasbourg/ Zone C : academies of Créteil, Montpellier, Paris, Toulouse and Versailles*

***Table S7.*** *Percentage reduction in contacts by each perturbation matrices for contacts at school*, at work and other contacts*

| Perturbation matrices | Percentage reduction in contacts at school | Percentage reduction in contacts at work | Percentage reduction in other contacts |
|---|---|---|---|
| S2 | -95% | | |
| S3 | -24% | | |
| S4 | -50% | | |
| S5 | -67% | | |
| S6 | -44% | | |
| O1 | | | -40% under 65<br>-70% over 65 |
| O2 | | | -90% under 65<br>-95% over 65 |
| O3 | | | -20% under 65<br>-50% over 65 |
| W1 | | -40% | |
| W2 | | -70% | |
| W3 | | -30% | |

*For contacts at school, we weighted the percentage reduction in contacts at school for each zone's vacations according to the population's size in each zone

***Table S8.*** *Application periods of perturbation matrices for contacts at school (S), at work (W) and other contacts (O)*

| Start | End | S | O | W | Start | End | S | O | W |
|---|---|---|---|---|---|---|---|---|---|
| 01/01/2020 | 12/03/2020 | 0 | 0 | 0 | 04/04/2021 | 06/04/2021 | 2 | 3 | 1 |
| 12/03/2020 | 14/03/2020 | 2 | 0 | 0 | 06/04/2021 | 26/04/2021 | 2 | 1 | 1 |
| 14/03/2020 | 17/03/2020 | 2 | 3 | 0 | 26/04/2021 | 03/05/2021 | 0 | 1 | 1 |
| 17/03/2020 | 02/06/2020 | 2 | 2 | 2 | 03/05/2021 | 12/05/2021 | 0 | 1 | 3 |
| 02/06/2020 | 22/06/2020 | 2 | 0 | 3 | 12/05/2021 | 16/05/2021 | 2 | 1 | 3 |
| 22/06/2020 | 04/07/2020 | 0 | 0 | 3 | 16/05/2021 | 18/05/2021 | 0 | 1 | 3 |
| 04/07/2020 | 01/09/2020 | 2 | 0 | 3 | 18/05/2021 | 20/06/2021 | 0 | 3 | 3 |
| 01/09/2020 | 17/10/2020 | 0 | 0 | 3 | 20/06/2021 | 06/07/2021 | 0 | 0 | 3 |
| 17/10/2020 | 30/10/2020 | 2 | 0 | 3 | 06/07/2021 | 01/09/2021 | 2 | 0 | 3 |
| 30/10/2020 | 01/11/2020 | 2 | 1 | 1 | 01/09/2021 | 23/10/2021 | 0 | 0 | 0 |
| 01/11/2020 | 15/12/2020 | 0 | 1 | 1 | 23/10/2021 | 08/11/2021 | 2 | 0 | 0 |
| 15/12/2020 | 19/12/2020 | 0 | 3 | 3 | 08/11/2021 | 18/12/2021 | 0 | 0 | 0 |
| 19/12/2020 | 04/01/2021 | 2 | 3 | 3 | 18/12/2021 | 01/01/2022 | 2 | 0 | 0 |
| 04/01/2021 | 06/02/2021 | 0 | 3 | 3 | 01/01/2022 | 07/02/2022 | 0 | 0 | 0 |
| 06/02/2021 | 13/02/2021 | 3 | 3 | 3 | 07/02/2022 | 14/02/2022 | 3 | 0 | 0 |
| 13/02/2021 | 22/02/2021 | 4 | 3 | 3 | 14/02/2022 | 21/02/2022 | 5 | 0 | 0 |
| 22/02/2021 | 01/03/2021 | 5 | 3 | 3 | 21/02/2022 | 28/02/2022 | 4 | 0 | 0 |
| 01/03/2021 | 07/03/2021 | 6 | 3 | 3 | 28/02/2022 | 07/03/2022 | 6 | 0 | 0 |
| 07/03/2021 | 04/04/2021 | 0 | 3 | 3 | 07/03/2022 | 01/04/2022 | 0 | 0 | 0 |

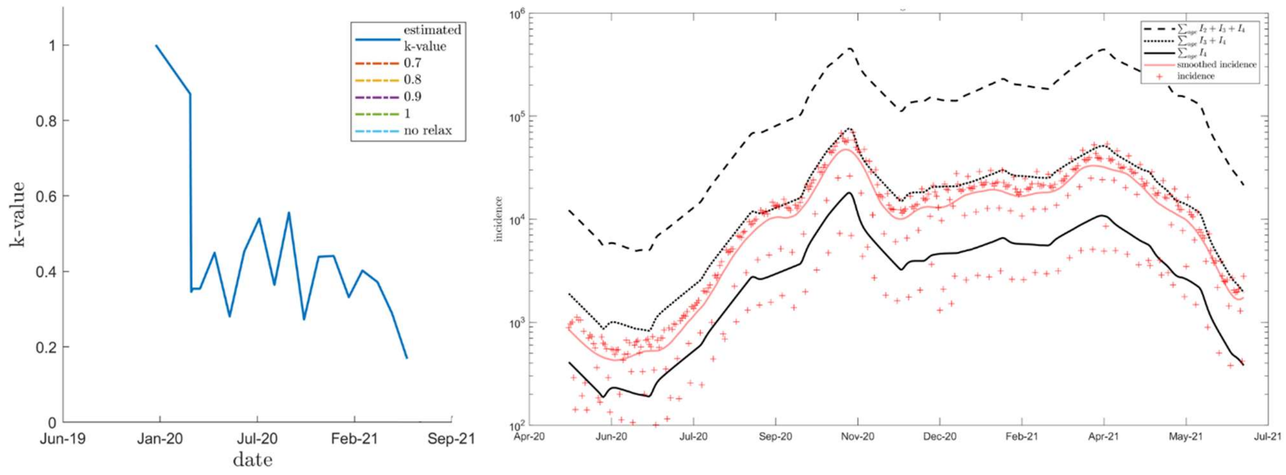


***(a)*** ***(b)***

***Figure S3.*** *Calibration's plot for the 3 years immunity duration analysis.* ***(a)*** *K-value estimation from May 13th, 2020 to July 1st, 2021.* ***(b)*** *Model fit (logarithmic scale) from May 13th, 2020 to July 1st, 2021 to reported COVID-19 cases from French surveillance database (SI-DEP-Système d'Informations de DEPistage)*

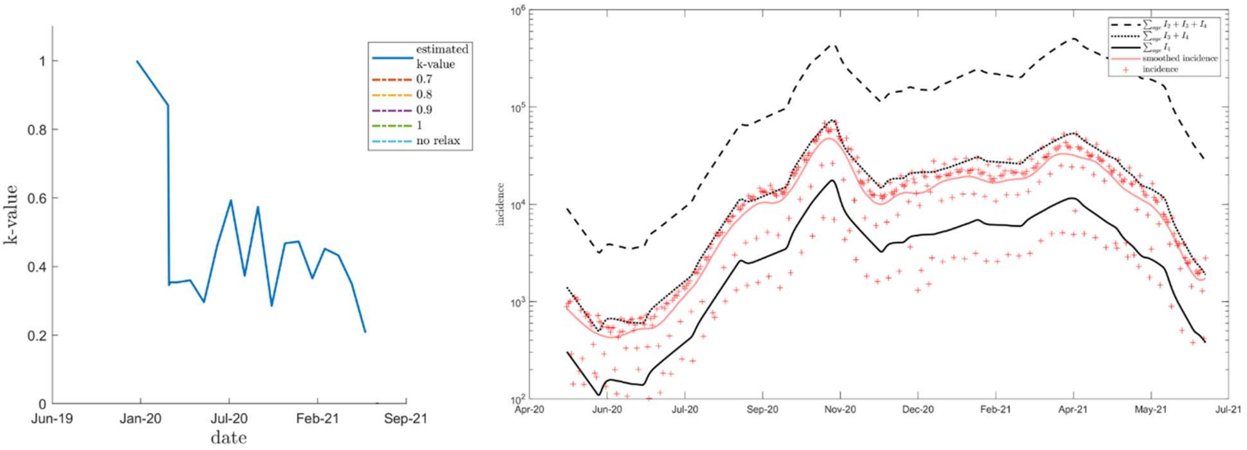


*(a)* *(b)*

***Figure S4.*** *Calibration's plot for the no waning immunity analysis.* ***(a)*** *K-value estimation from May 13th, 2020 to July 1st, 2021.* ***(b)*** *Model fit (logarithmic scale) from May 13th, 2020 to July 1st, 2021 to reported COVID-19 cases from French surveillance database (SI-DEP-Système d'Informations de DEPistage)*

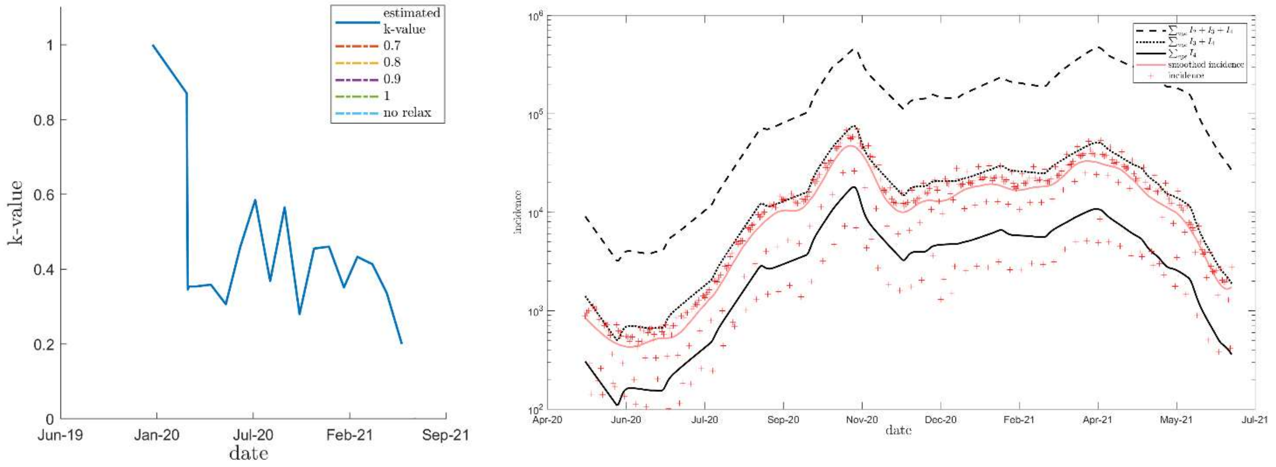


*(a)* *(b)*

***Figure S5.*** *Calibration's plot for the 9 years waning immunity analysis with a better vaccine efficacy.* ***(a)*** *K-value estimation from May 13th, 2020 to July 1st, 2021.* ***(b)*** *Model fit (logarithmic scale) from May 13th, 2020 to July 1st, 2021 to reported COVID-19 cases from French surveillance database (SI-DEP-Système d'Informations de DEPistage)*

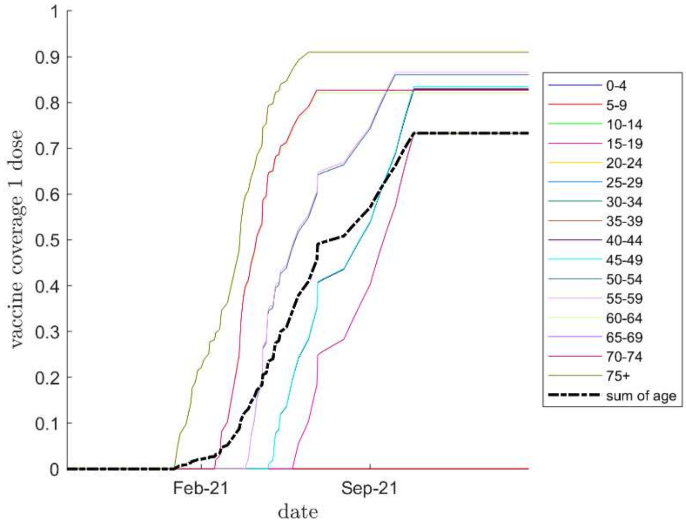


***Figure S6***. *Proportion of the population vaccinated with at least one dose by age class, taking into account vaccine hesitancy*

***Table S9***. *Proportion of the population vaccinated with at least one dose and vaccinated with 2 doses at the end of each month taking into account vaccine hesitancy*

| Month | Proportion vaccinated with 1 or 2 doses (%) | Proportion vaccinated with 2 doses (%) |
|---|---|---|
| January | 1.4 | 0.4 |
| February | 2.7 | 1.6 |
| March | 10.6 | 4.7 |
| April | 20.9 | 12.3 |
| May | 34.4 | 23.3 |
| June | 49.1 | 34.4 |
| July | 50.9 | 44.1 |
| August | 57.1 | 50.1 |
| September | 66.2 | 57.2 |
| October | 73.3 | 65.4 |
| November | 73.3 | 69.1 |
| December | 73.3 | 70.3 |

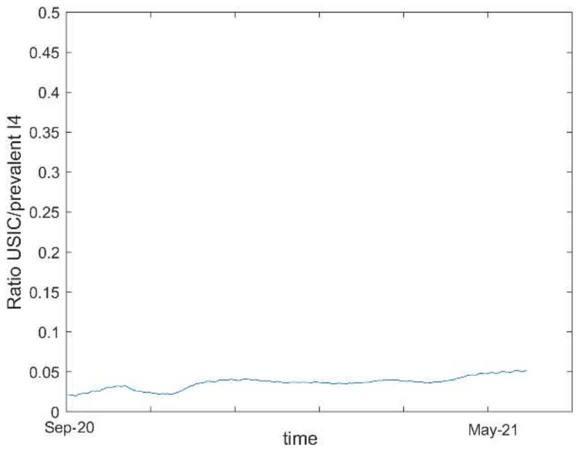


***Figure S7.*** *Ratio between intensive care units' (ICU) hospitalizations from SI-VIC database and prevalent $I_4$ cases from September 1st, 2020 to May 31th, 2021. SI-VIC = Système d'Information pour le suivi des VICtimes d'attentats et de situations sanitaires exceptionnelles.*

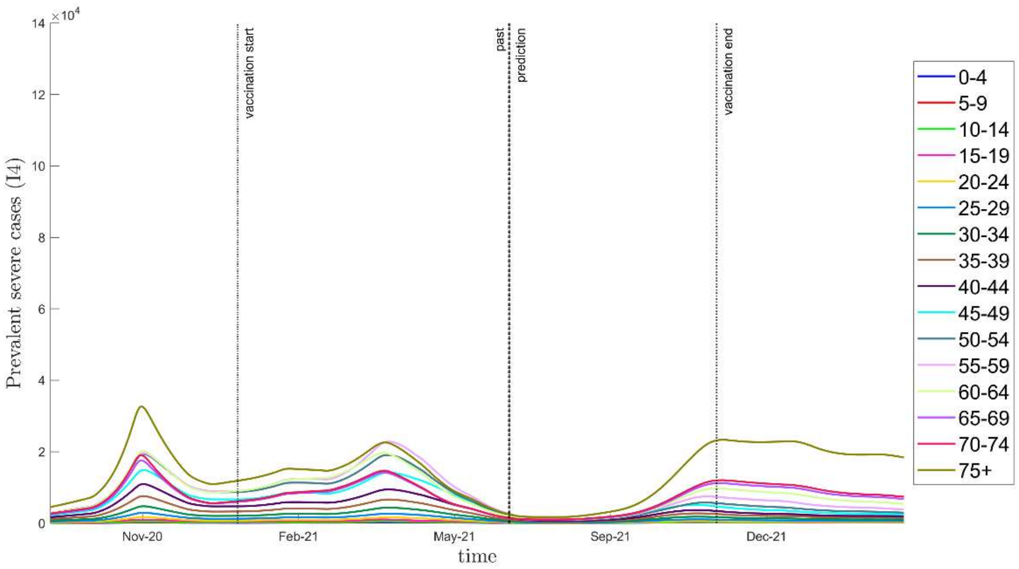


***Figure S8****. Prevalent severe cases $I_4$ by age class in the no barrier gesture relaxation scenario, assuming that immunity wanes in 9 years (main analysis)*

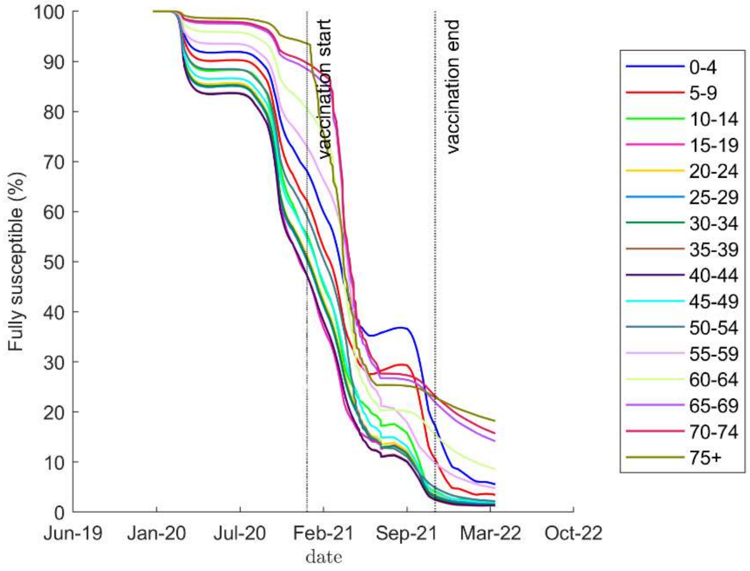


***Figure S9***. *Proportion of fully susceptible individuals ($S_1$) in each age class in the no barrier gesture relaxation scenario, assuming that immunity wanes in 9 years (main analysis)*

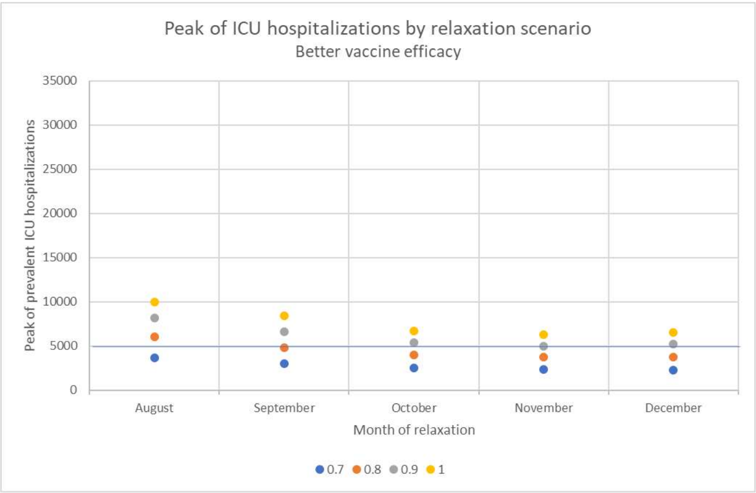


***Figure S10.*** *Peak of Intensive Care Units' hospitalizations (ICU) predicted in France from July 2021 following each relaxation scenario assuming a better vaccine efficacy. Relaxation scenarios: k-value (barrier gesture compliance index) raised either to 0.7, 0.8, 0.9 or 1 in August, September, October, November or December. Better vaccine efficacy = 70 and 90% vaccine efficacy against infections after respectively 1 and 2 doses.*

***Table S10****. Average variation of intensive care units' (ICU) hospitalizations peak predicted following each relaxation scenario assuming a better vaccine efficacy compared to the predictions obtained in the main analysis*

| Vaccine efficacy hypothesis | Average variation of ICU peak compared to main analysis* (%) |
|---|---|
| 70% and 90% vaccine efficacy against infections after respectively 1 and 2 doses | -41.0 |

*Average value of the peak of ICU hospitalizations predicted following each relaxation scenario (k-value elevated at 0.7, 0.8, 0.9 or 1 in August, September, October, November and December). *Main analysis = 50 and 80% vaccine efficacy against infections after respectively 1 and 2 doses.*

***Table S11****. Average variation of intensive care units' (ICU) hospitalizations peak predicted following each relaxation scenario for each hypothesis of immunity duration compared to the predictions obtained in the 9 years immunity duration analysis (main analysis)*

| Hypothesis of immunity duration | Average variation of ICU peak compared to main analysis* (%) |
|---|---|
| 3 years immunity duration | +86.1 |
| No waning immunity | -46.1 |

*Average value of the peak of ICU hospitalizations predicted following each relaxation scenario (k-value elevated at 0.7, 0.8, 0.9 or 1 in August, September, October, November and December)

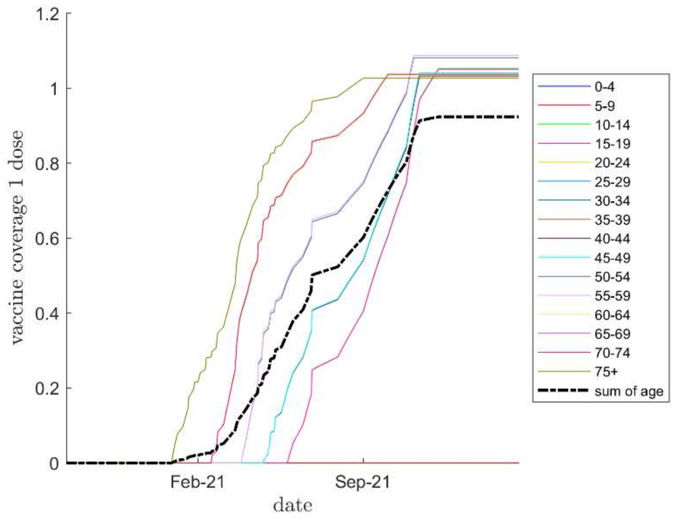


***Figure S11****. Proportion of the population vaccinated with at least one dose by age class assuming no vaccine hesitancy*

***Table S12***. *Proportion of the population vaccinated with at least one dose and vaccinated with 2 doses at the end of each month assuming no vaccine hesitancy*

| Month | Proportion vaccinated with 1 or 2 doses (%) | Proportion vaccinated with 2 doses (%) |
|---|---|---|
| January | 1.4 | 0.4 |
| February | 2.7 | 1.6 |
| March | 10.6 | 4.8 |
| April | 21.0 | 12.4 |
| May | 34.5 | 23.4 |
| June | 50.2 | 34.5 |
| July | 52.4 | 45.0 |
| August | 60.2 | 52.1 |
| September | 72.7 | 61.3 |
| October | 87.6 | 72.4 |
| November | 92.4 | 83.2 |
| December | 92.4 | 87.4 |

***Table S13.*** *Average variation of intensive care units' (ICU) hospitalizations peak predicted following each relaxation scenario assuming no vaccine hesitancy compared to the main analysis with vaccine hesitancy*

| Month of relaxation | Average variation of ICU peak compared to main analysis* (%) |
|---|---|
| August | -30.0 |
| September | -34.1 |
| October | -44.9 |
| November | -63.0 |
| December | -66.5 |

*Average value of the peak of ICU hospitalizations predicted following each relaxation scenario (k-value elevated at 0.7, 0.8, 0.9 or 1 for each month of relaxation)

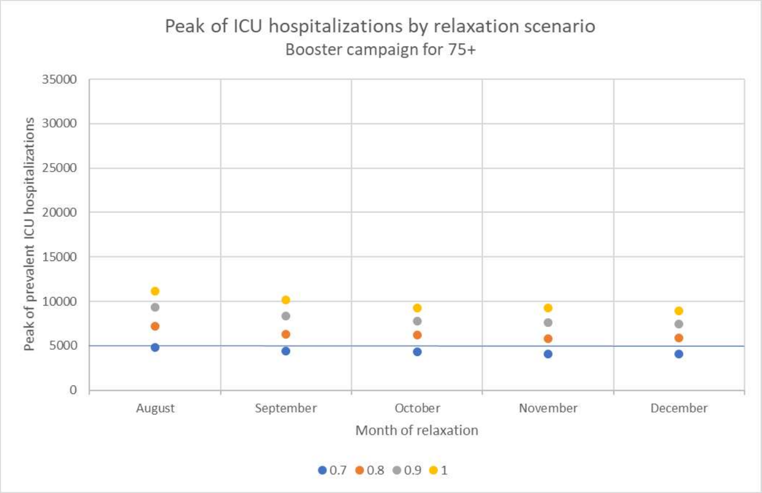


***Figure S12.*** *Peak of Intensive Care Units' hospitalizations (ICU) predicted in France from July 2021 following each relaxation scenario assuming a booster vaccination campaign on the September 1st, 2021 for the 75+. Relaxation scenarios: k-value (barrier gesture compliance index) raised either to 0.7, 0.8, 0.9 or 1 in August, September, October, November or December.*

***Table S14****. Average variation of intensive care units' (ICU) hospitalizations peak predicted following each relaxation scenario assuming a booster vaccination campaign on the September 1st, 2021 for the 75+ compared to the main analysis without booster*

| | Average variation of ICU peak compared to main analysis* (%) |
|---|---|
| Booster vaccination campaign for the 75+ | -16.3 |

*Average value of the peak of ICU hospitalizations predicted following each relaxation scenario (k-value elevated at 0.7, 0.8, 0.9 or 1 in August, September, October, November and December)
